# Temporal trends in the incidence of haemophagocytic lymphohistiocytosis: a nationwide cohort study from England 2003-2018

**DOI:** 10.1101/2022.03.30.22273090

**Authors:** Joe West, Peter Stilwell, Hanhua Liu, Lu Ban, Mary Bythell, Tim R Card, Peter Lanyon, Vasanta Nanduri, Judith Rankin, Mark Bishton, Colin J Crooks

**Affiliations:** Population and Lifespan Sciences, University of Nottingham; National Disease Registration Service, NHS Digital; NIHR Biomedical Research Centre, Nottingham University Hospitals NHS Trust; PPD Beijing; Watford General Hospital NHS Trust; Population Health Sciences Institute, Newcastle University; Translational Medical Sciences, University of Nottingham

**Keywords:** Haemophagocytic lymphohistiocytosis, incidence, epidemiology, rare disease, cohort study, inflammatory rheumatological disease, inflammatory bowel disease, haematology, malignancy

## Abstract

**Background:** Haemophagocytic lymphohistiocytosis (HLH) is rare, results in high mortality and is increasingly being diagnosed. Little is known about what is driving the apparent rise in the incidence of this disease.

**Methods:** Using national linked electronic health data from hospital admissions and death certification cases of HLH that were diagnosed in England between 1/1/2003 and 31/12/2018 were identified using a previously validated approach. We calculated incidence rates of diagnosed HLH per million population using mid-year population estimates by calendar year, age group, sex and associated comorbidity (haematological malignancy, inflammatory rheumatological or bowel diseases (IBD)) associated with the diagnosis of HLH. We modelled trends in incidence and the interactions between calendar year, age and associated comorbidity using Poisson regression.

**Findings:** There were 1674 people with HLH diagnosed in England between 2003 and 2018. The incidence rate quadrupled (Incidence Rate Ratio (IRR) 2018 compared to 2003: 3.88 95% Confidence Interval (CI) 2.91 to 5.28), increasing 11% annually (adjusted IRR 1.11 95% CI 1.09 to 1.12). There was a rising trend in all age groups except those aged less than 5 years. There was a transition across the age groups with greater increases in those aged 5 to 14 years of HLH associated with rheumatological disease/IBD compared to HLH associated with haematological malignancy, with similar increases in HLH associated with both co-morbidities for those 15-54, and greater increases in associated haematological malignancies for those 55 years and older.

**Interpretation:** The incidence of HLH in England has quadrupled between 2003 and 2018, increasing 11% annually. Substantial variation in the incidence occurred by age group and by HLH associated comorbidities with inflammatory rheumatological diseases or IBD associated HLH increasing more among the young and middle age groups, whereas in older age groups the largest increase was seen with haematological malignancy-associated HLH.

**Evidence before this study:** There is a paucity of population-based data on the epidemiology of HLH worldwide. The available evidence relies mostly upon a collection of cases series published in The Lancet in 2014 which described 2197 cases of HLH in adults reported in the literature to that point. Almost all of these were from tertiary referral specialist centres and/or described in small case series. The incidence of HLH has only been described in a few reports – and mainly this has focused on children with primary HLH. No previous studies have been large enough to examine trends in incidence by age, sex, underlying risk factors and calendar time.

**Added value of this study:** This study quantifies the incidence of diagnosed HLH for the first time in a nationwide manner for all age groups. It reports on 1674 patients with HLH from England and shows that there is substantial variation in the incidence by age group, associated disease and calendar time. The results imply reasons for the increase in HLH could be related to the increasing occurrence of haematological cancer, inflammatory rheumatological or bowel diseases and the treatments given for these conditions. This study has been carried out in partnership with the National Congenital Anomalies and Rare Diseases Registration Service and the methodology described can in future be applied to many rare diseases that as yet lack a way of quantifying crucial epidemiological metrics.

**Implications of all the available evidence:** The incidence of HLH is rising rapidly in people older than 5 years of age. This could be due to an increase in the biologic, immunomodulation or immunosuppressive therapy being used in people with haematological cancer and inflammatory rheumatological and bowel diseases. Further work should focus on how to minimise the risk of HLH occurring, or to improve treatment of this often fatal disease among those who need treatment for an associated comorbidity.

## Introduction

Understanding the causes and burden of disease for the purposes of planning optimal health care requires contemporary population-level estimates of disease incidence^1^. Although attempts to do so have been made, improving our ability to quantify and describe the occurrence of rare diseases remains a worldwide challenge^2^. Precisely measuring the occurrence of a rare disease in a population is difficult as the population needs to be large enough to adequately quantify temporal trends in incidence and variation by important socicodemographic and clinical factors, whilst minimising the bias inherent in case reports or small cohorts compiled in specialist centres^3^. Haemophagocytic lymphohistiocytosis (HLH) is a rare clinical syndrome characterised by fever, hyper-inflammation, organ dysfunction, cytopenias and haemophagocytosis^4^ and, as it can be an inherited or acquired disorder affecting all ages and has several risk factors, is an appropriate exemplar. HLH is associated with high mortality rates in all age groups^4^ particularly those with underlying malignancy^5^. However, the incidence of HLH on a population level has rarely been quantified^6-8^ with only one previous report containing information on both children and adults from the same population^9^. That report indicated that the incidence of HLH appears to be increasing over time but it was unable to explore the reasons why as despite being the largest incidence study in the literature it was still too small to examine how trends over time varied by age, sex and the underlying diseases associated with HLH. A combination of the increasing incidence of non-Hodgkin lymphoma^10^, increasing use of biologic therapies or immune-suppressants in autoimmune inflammatory diseases^11,12^ increasing age of first exposure to the Epstein-Barr virus (EBV) or increasing clinical awareness could be contributing to the observed rise in HLH^13^.

To examine the temporal trends in incidence of HLH and explore the relationships with demographic and clinical characteristics, we have carried out a nationwide study in England in partnership with the National Disease Registration Service (NDRS).

## Methods

### Data sources

We used linked electronic health records from English Hospital Episode Statistics (HES)^14^, the National Cancer Registration Dataset (NCRD)^15^ and Office for National Statistics (ONS) death certification data. In brief, HES data are collected routinely for all National Health Service (NHS) related hospital admissions in England and contain sociodemographic and clinical information, the latter coded with International Classification of Diseases, 10^th^ revision (ICD-10) and Office of Population, Censuses and Surveys Classification of Surgical Operations and Procedures version 4 (OPCS-4), NCRD contains all cancers registered in England and their associated histological and staging information and ONS death certification data contains the contents of the death certificate coded for underlying cause of death (with ICD-10), including any associated free text.

### Study population and case identification

Patients diagnosed with HLH were identified using our previously validated approach^16^. They included HLH patients of all ages who were admitted to hospital or died between 1 January, 2003 and 31 December, 2018 in England. In brief, we included people who had an admission coded with ICD-10 codes for HLH – D76.1 (Haemophagocytic lymphohistiocytosis) or D76.2 (Haemophagocytic syndrome) or a death coded with D76.1, D76.2 or D76.3 (Other histiocytosis syndromes) as long as in the latter situation there was confirmatory free text on the death certificate indicating HLH. The date of diagnosis was taken as the first day of the hospital admission in which HLH was coded or, if identified only by death registration, the date of death.

### HLH associated characteristics and associated comorbidities

For all patients we extracted information on age and sex at diagnosis and determined HLH associated comorbidities. The presence or absence of comorbidities was identified from all available HES and NCRD records prior to the diagnosis of HLH and up to three months after diagnosis. We defined haematological cancer, non-haematological cancer (excluding non-melanoma skin cancer), inflammatory rheumatological disease, inflammatory bowel disease (IBD), herpes viruses (cytomegalovirus (CMV), EBV and varicella zoster virus (VZV)) and human immunodeficiency virus/acquired immune deficiency syndrome (HIV/AIDS) using ICD-10 codes (supplementary tables S1, S2 and S3). For inflammatory rheumatological diseases we assigned subcategories of diagnosis (supplementary data table S1) i.e. systemic juvenile idiopathic arthritis, systemic lupus erythematosus (SLE) etc. If more than one subcategory of inflammatory rheumatological disease was identified, the patient was assigned to the diagnosis group that was recorded closest in time to the HLH diagnosis. For haematological malignancy, if two malignancies were recorded patients were assigned to both subcategories. For the purposes of later analyses where there was overlap of non-infectious associated comorbidities we classified patients with the following mutually exclusive hierarchy: haematological malignancy, rheumatological disease/IBD, non-haematological malignancy and none of these recorded. For patients where more than one malignancy was recorded a panel of authors (JW, CJC, MBishton, PS) considered the different diagnoses and temporal relationship to the diagnosis of HLH for each case. For the purposes of hierarchical coding, the malignancy assigned was the diagnosis that occurred within the period 3 years prior to HLH and up to one month after the HLH diagnosis. If the malignancies were prior to 3 years before the HLH diagnosis, the haematological malignancy was assigned. As there was not access to serological tests for the cohort, the type (acute, chronic, reactivation) of viral illness could not be ascertained and so we have not included these in the “associated diseases hierarchy”.

### Statistical analysis

Numbers and frequencies of the patient’s characteristics were calculated and an assessment of those cases with more than one HLH associated comorbidity was carried out via cross tabulation and plotting a Venn diagram. The incidence rate (equivalent to the frequency of clinical diagnosis) of HLH was calculated by summing the number of cases (including those diagnosed via death certificate only) within age group, sex and calendar year strata and dividing by the summed person years within each stratification variable based on the ONS mid-year population estimates. Incidence rates of HLH cases with associated comorbidities were calculated by including only the relevant cases (only for the chronic, non-infectious morbidities) over the whole population denominator. Incidence rates were then presented by calendar year, age group, sex and associated comorbidity with 95% confidence intervals around the estimates derived via a Poisson distribution. A Poisson regression model was fitted to estimate the age group and sex adjusted incidence rate ratios (IRR) associated with calendar year (fitted as a categorical variable). To estimate the annual % change in incidence a further model was fitted using calendar year as a linear term adjusted for age group and sex. This model was compared to the model with calendar year as a categorical variable with a likelihood ratio test to assess evidence of departure from a linear trend. To assess temporal trends in incidence within age group, by the various HLH associated comorbidity groups and by age group and associated comorbidity, we plotted incidence rates over time within age groups, by each HLH associated comorbidity group and both together. For ease of display in the latter case we plotted the moving average of the incidence rate using a locally estimated scatterplot smoothing curve (LOESS). To quantify these time trends we fitted a model containing a 2-way interaction between age group and calendar year as a linear term, and separately associated comorbidity and calendar year. We modelled changes in associated comorbidity within age groups by fitting a 2-way interaction within age stratified models. Finally, we computed stratum-specific adjusted IRRs and 95% CIs for each age group, associated comorbidity and jointly to estimate the annual increase in incidence over the study period, with 2003 as the reference category.

Data for this study were collected and analysed under the National Disease Registries Directions 2021, made in accordance with sections 254(1) and 254(6) of the 2012 Health and Social Care Act. Ethical approval for this study was not required per the definition of research according to the UK Policy Framework for Health and Social Care Research. The data are available to those that have the legal basis to access it, either through the Data Access Request Service (https://digital.nhs.uk/services/data-access-request-service-dars) or partnership with NDRS. The protocol was approved by the joint NDRS project board reference PPF1920_027. Study findings are reported in accordance with the REporting of studies Conducted using Observational Routinely-collected health Data (RECORD) recommendations^17^. We used R (version 4.1.2)^18^ for the data management and statistical analyses.

## Results

In the 16-year study period 2003 to 2018, we identified 1674 patients with an incident diagnosis of HLH (supplementary figure 1 – study flow diagram) via our validated approach^16^. The majority, 1091 (65%), were identified through an admission to hospital coded with either ICD10 D76.1 or D76.2 in HES, while a further 183 (11%) were identified at death using the same codes, or a D76.3 code with confirmatory free text, used anywhere on their death certificates. The remainder were identified in both HES and ONS mortality data. Of the whole cohort 551 (33%) had a record in the NCRD indicating a registered cancer (excluding non-melanoma skin cancer) prior to or up to three months after their HLH diagnosis. Of the whole cohort 534 (32%) had a recorded associated non-malignant comorbidity (inflammatory rheumatological disease/IBD/viral) and the majority of chronic autoimmune comorbidities were recorded in hospital admissions prior to the admission in which the HLH diagnosis occurred (92%). The characteristics of the cases are described in table 1 and overlap of associated comorbidities in figure 1. Almost 70% were aged ≥15 years, over half (56%) were male and just over half (51%) of people had at least one of the associated comorbidities recorded in their records. Of these diseases, the commonest single disease entities were lymphoma, leukaemia, systemic juvenile idiopathic arthritis (SJIA), IBD, vasculitis and systemic lupus erythematosus (SLE). There was evidence of clinically diagnosed EBV, CMV, VZV infection or HIV/AIDS in 7.6%, 5.7%, 1.2% and 0.6% respectively. 107 (6.4%) patients had a record of an allogeneic peripheral blood stem cell transplant and 27 (1.6%) had a record of an autologous peripheral blood stem cell transplant. The majority of these stem cell transplants in children occurred after the diagnosis of HLH (82%) whereas for adults 50% occurred after the HLH diagnosis. In terms of associated comorbidities less than 5% of cases had more than one recorded. The largest overlap in associated comorbidities was between haematological malignancies and rheumatological disease/IBD (figure 1).

**Table 1.** Characteristics of the HLH cohort with incidence rates per million population

| Characteristic |  | n | % | Rate* | CI (95%)† |
| --- | --- | --- | --- | --- | --- |
| Sex | Female | 729 | 43.55 | 1.70 | (1.57 to 1.82) |
|  | Male | 945 | 56.45 | 2.27 | (2.13 to 2.42) |
| Calendar year | 2003 | 53 | 3.17 | 1.06 | (0.8 to 1.39) |
|  | 2004 | 42 | 2.51 | 0.84 | (0.6 to 1.13) |
|  | 2005 | 44 | 2.63 | 0.87 | (0.63 to 1.17) |
|  | 2006 | 55 | 3.29 | 1.08 | (0.81 to 1.4) |
|  | 2007 | 63 | 3.76 | 1.23 | (0.94 to 1.57) |
|  | 2008 | 63 | 3.76 | 1.22 | (0.93 to 1.56) |
|  | 2009 | 77 | 4.60 | 1.48 | (1.16 to 1.84) |
|  | 2010 | 86 | 5.14 | 1.63 | (1.31 to 2.02) |
|  | 2011 | 95 | 5.68 | 1.79 | (1.45 to 2.19) |
|  | 2012 | 120 | 7.17 | 2.24 | (1.86 to 2.68) |
|  | 2013 | 122 | 7.29 | 2.26 | (1.88 to 2.7) |
|  | 2014 | 141 | 8.42 | 2.60 | (2.19 to 3.06) |
|  | 2015 | 142 | 8.48 | 2.59 | (2.18 to 3.05) |
|  | 2016 | 141 | 8.42 | 2.55 | (2.15 to 3.01) |
|  | 2017 | 194 | 11.59 | 3.49 | (3.01 to 4.01) |
|  | 2018 | 236 | 14.10 | 4.22 | (3.70 to 4.79) |
| Age | 0-4 | 324 | 19.35 | 6.30 | (5.63 to 7.02) |
|  | 5-14 | 196 | 11.71 | 1.96 | (1.69 to 2.25) |
|  | 15-54 | 577 | 34.47 | 1.27 | (1.17 to 1.38) |
|  | 55+ | 577 | 34.47 | 2.41 | (2.22 to 2.62) |
| Chronic conditions | Inflammatory bowel disease | 70 | 4.18 | 0.08 | (0.06 to 0.1) |

| Characteristic |  | n | % | Rate* | CI (95%)<sup>†</sup> |
| --- | --- | --- | --- | --- | --- |
|  | Inflammatory rheumatological disease | 322 | 19.24 | 0.38 | (0.34 to 0.42) |
|  | Systemic juvenile idiopathic arthritis | 82 | 4.90 | 0.10 | (0.08 to 0.12) |
|  | Systemic lupus erythematosus | 48 | 2.87 | 0.06 | (0.04 to 0.08) |
|  | Adult-onset Still's disease | 31 | 1.85 | 0.04 | (0.02 to 0.05) |
|  | Rheumatoid arthritis | 43 | 2.57 | 0.05 | (0.04 to 0.07) |
|  | Vasculitis | 66 | 3.94 | 0.08 | (0.06 to 0.10) |
|  | Other inflammatory arthritis | 10 | 0.60 | 0.01 | (0.01 to 0.02) |
|  | Other connective tissue diseases | 42 | 2.51 | 0.05 | (0.04 to 0.07) |
| Malignancies | Any lymphoma condition<sup>‡</sup> | 279 | 16.67 | 0.33 | (0.29 to 0.37) |
|  | T-cell lymphoma | 84 | 5.02 | 0.10 | (0.08 to 0.12) |
|  | B-cell lymphoma | 121 | 7.23 | 0.14 | (0.12 to 0.17) |
|  | Hodgkin lymphoma | 45 | 2.69 | 0.05 | (0.04 to 0.07) |
|  | Lymphoma Not Otherwise Specified (NOS) | 32 | 1.91 | 0.04 | (0.03 to 0.05) |
|  | Leukaemia | 131 | 7.83 | 0.15 | (0.13 to 0.18) |
|  | Other haematological histiocytic / Myelodysplastic / Malignancy / Unspecified | 74 | 4.42 | 0.09 | (0.07 to 0.11) |
|  | Non haematological malignancy excluding non melanoma skin cancer | 112 | 6.69 | 0.13 | (0.11 to 0.16) |
| Hierarchical chronic conditions | Haematological malignancy | 467 | 27.90 | 0.55 | (0.50 to 0.60) |
|  | Rheumatological disease/IBD | 323 | 19.30 | 0.38 | (0.34 to 0.43) |
|  | Non-haematological malignancy excluding non-melanoma skin cancer | 75 | 4.48 | 0.09 | (0.07 to 0.11) |
|  | None of the above | 809 | 48.33 | 0.96 | (0.89 to 1.02) |

\* Rate per million population
<sup>†</sup> 95% Confidence interval
<sup>‡</sup> Multiple lymphoma conditions allowed

**Figure 1.**
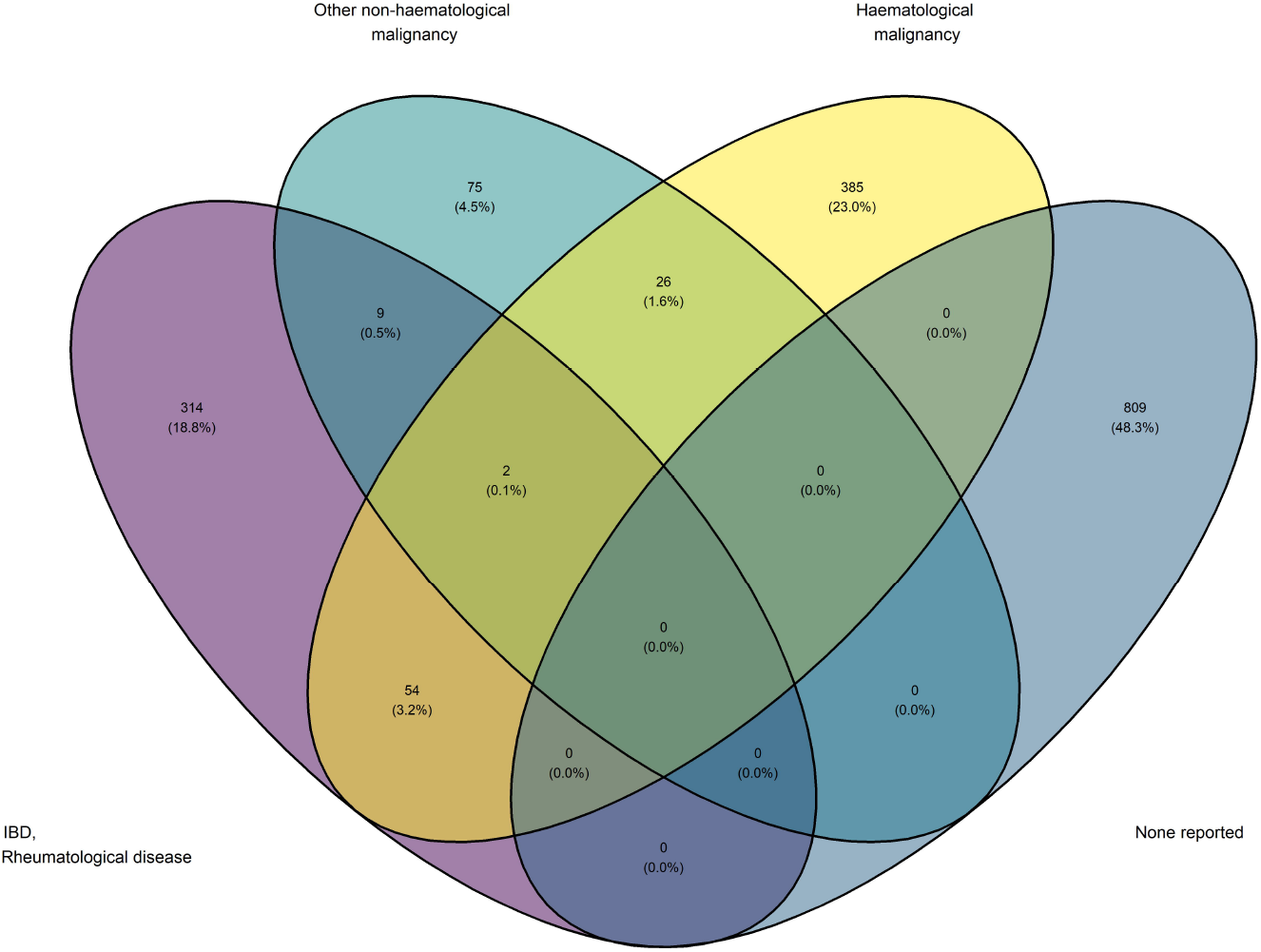
Venn diagram to show the overlap between of comorbidities within the whole HLH cohort

Reported crude incidence rates of HLH increased during the study period from around 1 per million person years in 2003 to around 4 per million in the 2018 (table 1) equating to an age and sex adjusted 4-fold (2018 compare to 2003: IRR 3.88 95% CI 2.91 to 5.28) increase in the reported incidence of HLH over the whole study period (supplementary table S4). Across the whole study period, the estimated year-on-year relative increase (calendar year fitted as a continuous variable), assuming a linear trend, of HLH incidence was 11% (IRR 1.11 95% CI 1.09 to 1.12). When compared to the model with calendar year as a categorical variable, there was no evidence of departure from a linear trend (p=0.32). This trend over time varied substantially by age (figure 2, p value for interaction <0.0001) such that there was no change in incidence over the study period in the under 5 year olds (IRR per year 1.00 95% CI 0.97 to 1.04) whereas there was an increasing rate in incidence over time in the 5-14, 15-54 and 55+ age groups (table 2).

**Figure 2.**
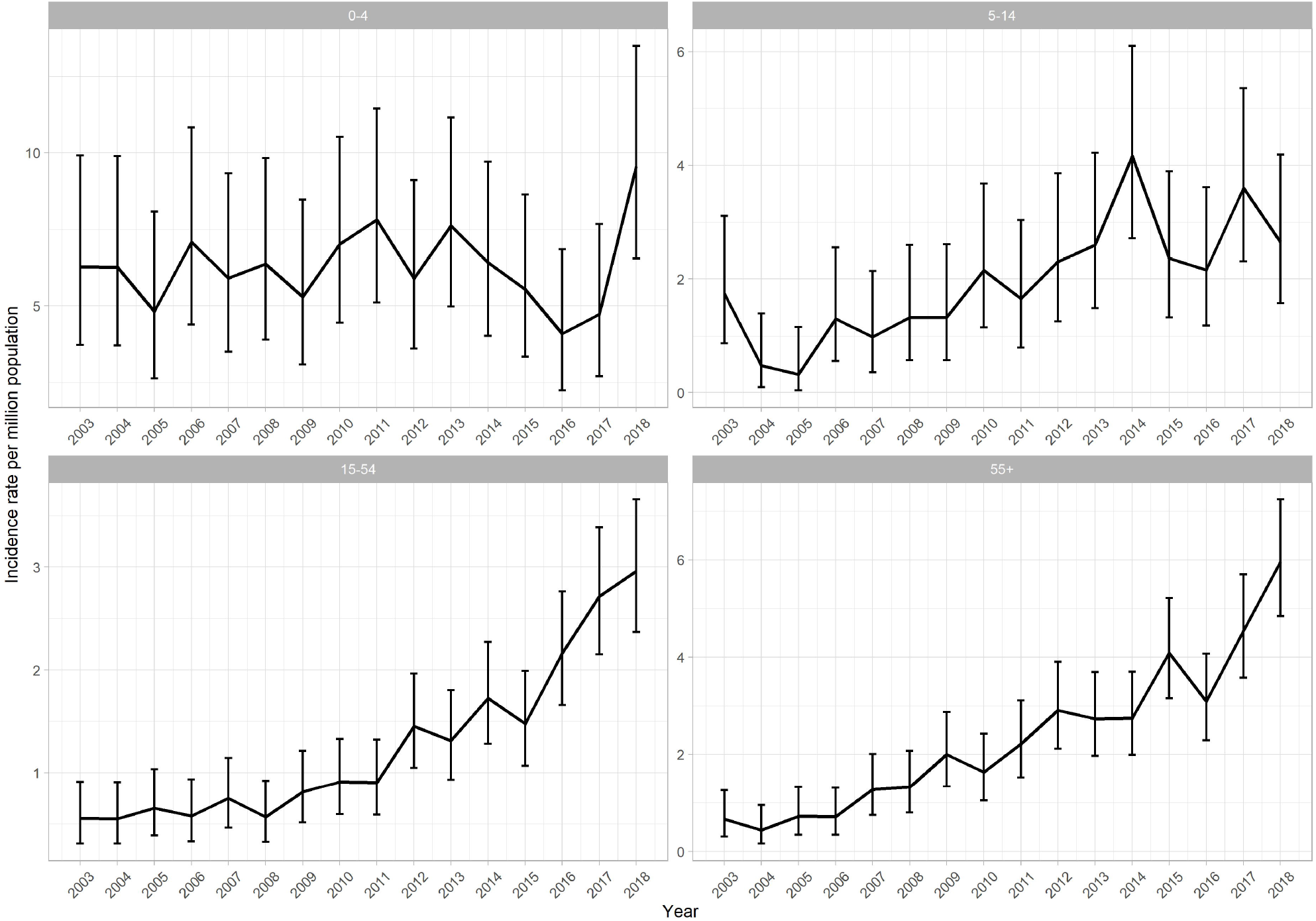
Incidence (95% CI) of HLH per million population over calendar time (2003-2018) and by age group

**Table 2.** Incidence rate ratios including the interaction between age group and calendar year

| Age group | IRR<sup>1</sup> | CI (95%)<sup>2</sup> |
| --- | --- | --- |
| 0-4 | 1.00 | (0.97 to 1.04) |
| 5-14 | 1.09 | (1.05 to 1.14) |
| 15-54 | 1.14 | (1.11 to 1.16) |
| 55+ | 1.16 | (1.13 to 1.19) |

<sup>1</sup> IRR = Incidence Rate Ratio: this represents the relative annual change in incidence of HLH (1.00 represents no change) for each age group, <sup>2</sup> CI = Confidence Interval

There was variation in the incidence rate of HLH over calendar time when stratified by the recorded presence of an associated comorbidity (figure 3, table 3, p value for interaction <0.0001) such that it was apparent that diagnoses of HLH with either haematological malignancy (IRR per year 1.17 95% CI 1.14 to 1.21), or rheumatological disease/IBD (IRR per year 1.20 95% CI 1.15 to 1.24) increased almost 20% per year. By contrast those with no recorded comorbidity (IRR per year 1.05 95% CI 1.03 to 1.07) or cases associated with non-haematological malignancy (IRR per year 1.12 95% CI 1.05 to 1.20) rose less markedly.

**Figure 3.**
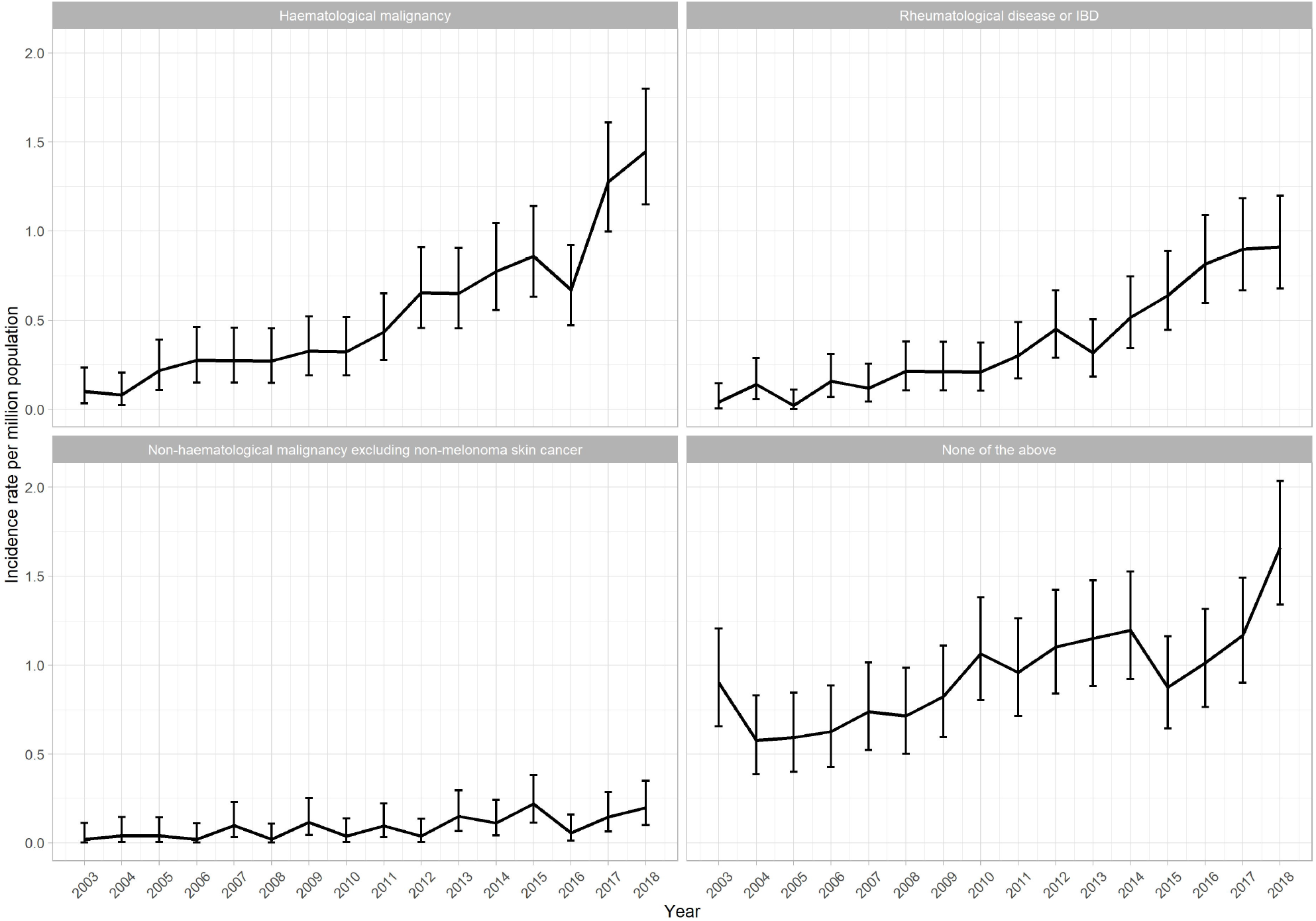
Incidence (95% CI) of HLH per million population over calendar time by associated comorbidity

**Table 3.**
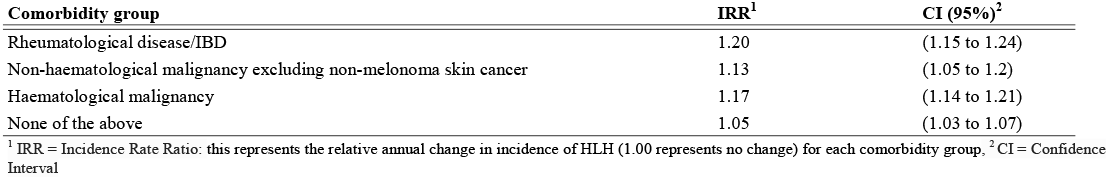
Incidence rate ratios including the interaction between comorbidity group and calendar year

Finally, we examined the effect on the temporal trends of the interaction between age group and associated comorbidities (figure 4, table 4, supplementary figure 2). This showed that there was a transition across the age groups with increases in incidence in those aged 5-14 years of HLH associated with rheumatological disease/IBD (IRR per year 1.12 95% CI 1.04 to 1.22) showing a 12% annual increase. When compared to HLH associated with haematological malignancy the 5-14 year olds had only a 4% annual increase (IRR per year 1.04 95% CI 0.94 to 1.14). In the 15-54 year age group, there were similar annual increases in HLH associated with both haematological malignancy and rheumatological disease/IBD (17% and 20% respectively), and for those 55 years and older there were increases in associated haematological malignancies (∼18% annual increase).

**Figure 4.**
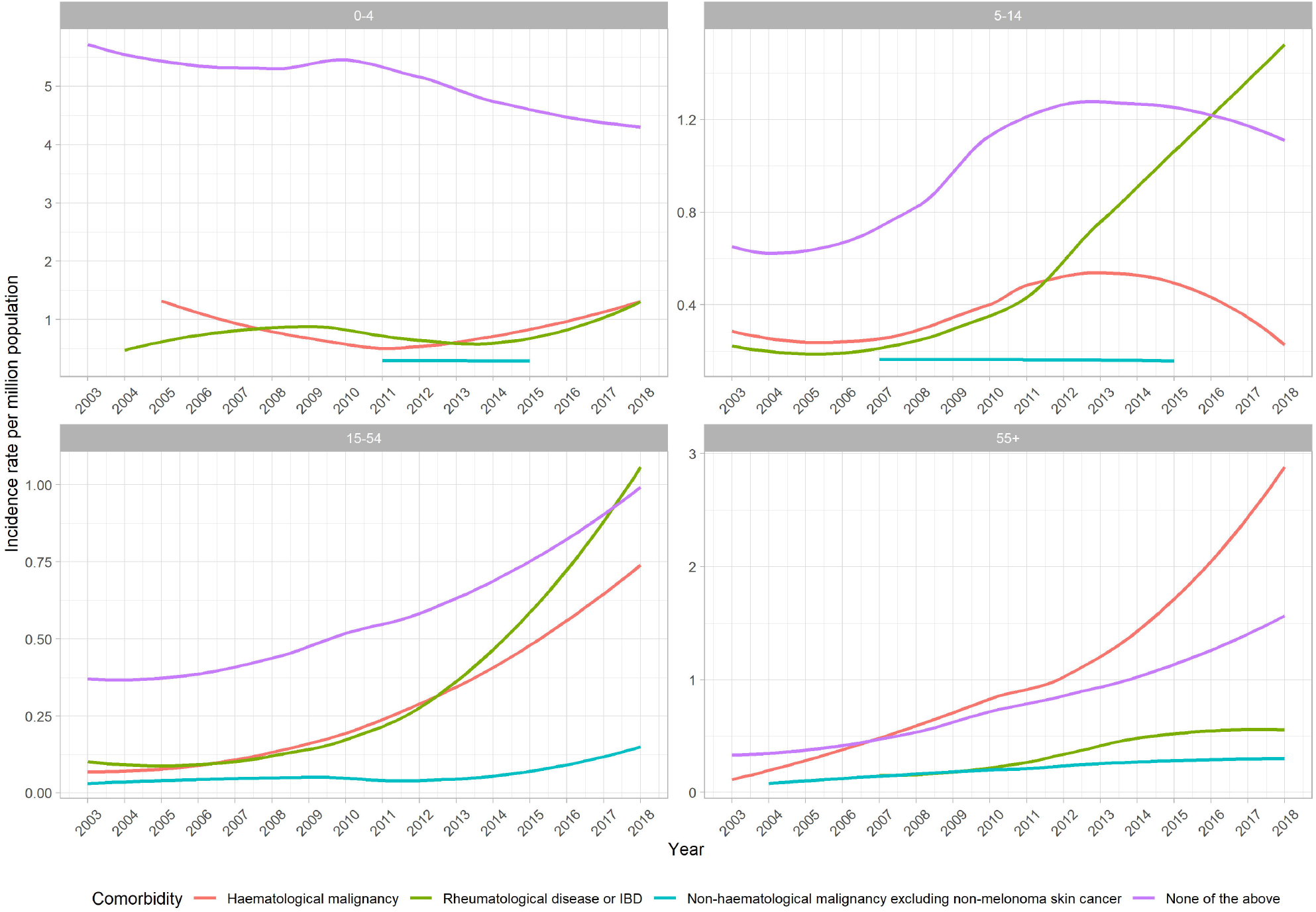
Incidence (LOESS) of HLH per million population over calendar time by age group and comorbidity

**Table 4.**
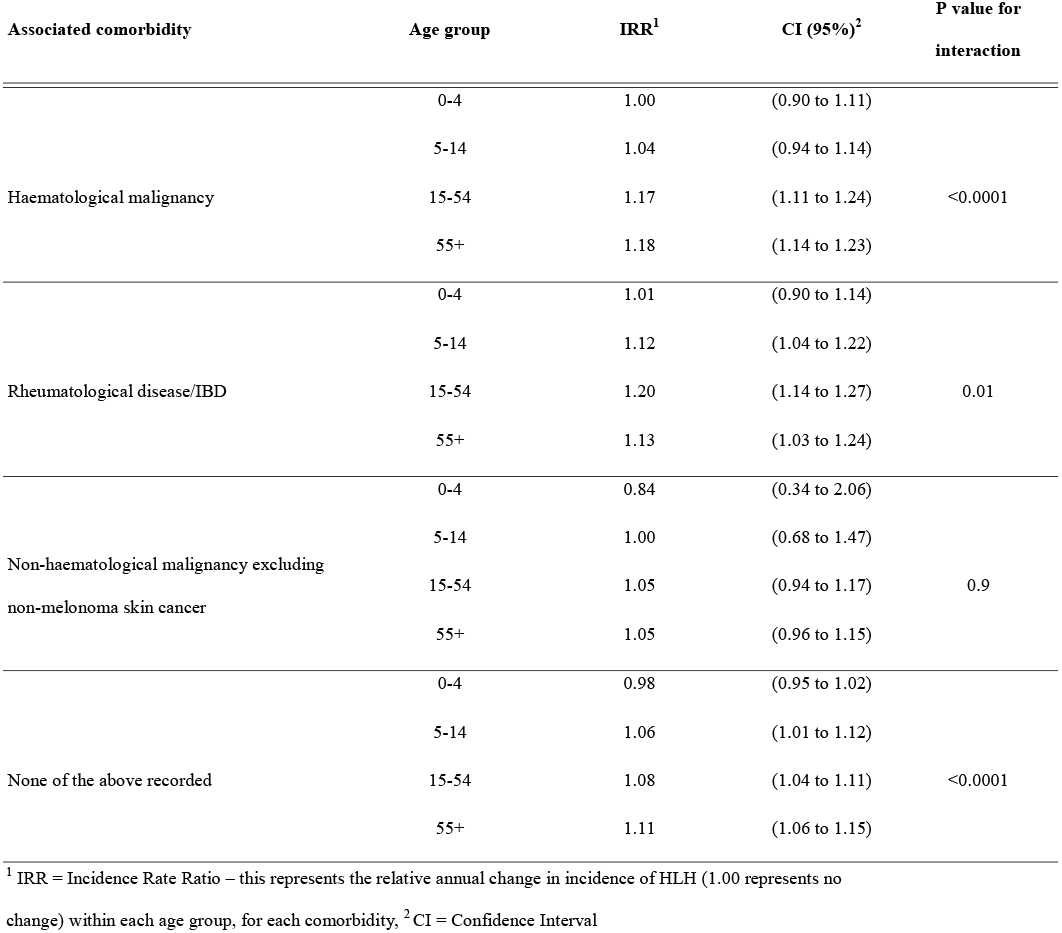
Incidence rate ratios including the interaction between age group, calendar year and associated comorbidity

| Associated comorbidity | Age group | IRR<sup>1</sup> | CI (95%)<sup>2</sup> | P value for interaction |
| --- | --- | --- | --- | --- |
| Haematological malignancy | 0-4 | 1.00 | (0.90 to 1.11) | <0.0001 |
|  | 5-14 | 1.04 | (0.94 to 1.14) |  |
|  | 15-54 | 1.17 | (1.11 to 1.24) |  |
|  | 55+ | 1.18 | (1.14 to 1.23) |  |
| Rheumatological disease/IBD | 0-4 | 1.01 | (0.90 to 1.14) | 0.01 |
|  | 5-14 | 1.12 | (1.04 to 1.22) |  |
|  | 15-54 | 1.20 | (1.14 to 1.27) |  |
|  | 55+ | 1.13 | (1.03 to 1.24) |  |
| Non-haematological malignancy excluding non-melanoma skin cancer | 0-4 | 0.84 | (0.34 to 2.06) | 0.9 |
|  | 5-14 | 1.00 | (0.68 to 1.47) |  |
|  | 15-54 | 1.05 | (0.94 to 1.17) |  |
|  | 55+ | 1.05 | (0.96 to 1.15) |  |
| None of the above recorded | 0-4 | 0.98 | (0.95 to 1.02) | <0.0001 |
|  | 5-14 | 1.06 | (1.01 to 1.12) |  |
|  | 15-54 | 1.08 | (1.04 to 1.11) |  |
|  | 55+ | 1.11 | (1.06 to 1.15) |  |

<sup>1</sup> IRR = Incidence Rate Ratio – this represents the relative annual change in incidence of HLH (1.00 represents no change) within each age group, for each comorbidity, <sup>2</sup> CI = Confidence Interval

## Discussion

The incidence of diagnosed HLH in England increased 11% year on year between 2003 and 2018 resulting in a 4-fold increase over the 16-year study period. Substantial variation in the incidence occurred by age groups, with no increase over time in those under 5 year olds contrasting with a 9% annual increase in 5-14 year olds, a 14% annual increase in 15-54 year olds and a 16% annual increase in those aged 55 and over. Furthermore, we observed increases in diagnoses of HLH associated with inflammatory rheumatological disease/IBD-and haematological malignancy-associated HLH over time, which also varied by age. Among the young and middle age groups there were increases in both rheumatological disease/IBD- and haematological malignancy-associated HLH, whereas in older age groups the increase was seen mainly with haematological malignancy-associated HLH. These findings imply that the temporal increase in HLH we have observed is being driven more in younger people by changes in autoimmune disease and its treatment, for example biologics and immunosuppressants, and more in older people by the known increase in incidence of haematological cancer, particularly non-Hodgkin lymphoma, during the study period^10^. Part of the increase in the rate of diagnosed HLH is likely to be due to the groups of clinicians involved in managing the associated comorbidities increasingly recognising HLH during the study period.

Our case definition of HLH is based on a published validation exercise we carried out in five English NHS Trusts (hospitals) that showed a positive predictive value for a diagnosis of HLH of 89.0% (95% CI 80.2-94.9%)^19^. In a systematic review of validation studies of other diseases, HES recording has been shown to be accurate for the purposes of research in this manner^20^. Two other studies in France^21^ and Chicago, USA^22^ have used a similar algorithm to identify cases of HLH and NHS Digital, who oversee coding within the NHS in England, confirmed that no changes have occurred in the procedural use of D76.1, D76.2 and D76.3 during the study period. We are therefore reasonably confident that the cases we have included represent true diagnoses of HLH, and that these diagnoses are underpinned by clinical use of diagnostic scoring systems such as the “HScore”^23^ or the “HLH-2004 diagnostic criteria”^24,25^. Our study design does mean that there will have been under-ascertainment of HLH as it is recognised to be a difficult disease to diagnose^4^ and therefore our estimates of incidence are likely to be, in general, underestimates as cases of HLH that occurred but were not clinically diagnosed as such in the hospital setting would not have been included in our study.

The results in the current study confirm those found in our recent report that utilised a smaller sample of the English population (via the Clinical Practice Research Datalink^26^ (CPRD)) over a similar time period^9^ – a rising incidence rate and variation by age. The current report encompasses the whole of England and therefore is not subject to selection bias with a sample size eight times bigger than our original study. Due to this size we were able to investigate temporal trends in incidence by age group and associated comorbidities, for all age groups, in a manner that no prior study of HLH has been able to do. Indeed, our use of a national population-based cohort means that the number of patients we report with HLH and haematological cancer – 467 – is far larger than any other single study in the literature to date^4^. The characteristics of our cohort are, as might be expected, similar to our CPRD study^9^ but also to the most relevant, recent, comparable studies of HLH from France^21^ Germany^27^, China^28,29^, Sweden^6^ and the USA^30,31^ in terms of age and sex distributions and also the proportions with haematological malignancy and rheumatological diseases.

Reasons why we have observed such a marked increase in the incidence rate of HLH in England over time and the variation by both age and associated comorbidities require some explanation. The increasing rates of diagnosis over time would, in part, be due to increasing frequency of recognition and diagnosis of HLH – essentially an increase in ascertainment. Notably, though, the rises in incidence over time were not observed in the under 5 year age group which will be, in the majority, “primary HLH” due to inherited risk factors. If we accept that not all of the increase in frequency of diagnosis in those aged 5 or older is due to ascertainment, then we can speculate on the alternative causes of the temporal trends. During the study period there has been a steady increase in the incidence of haematological malignancy, particularly non-Hodgkin lymphoma^10^, and in the prevalence of some inflammatory rheumatic diseases^32-34^ and IBD^35^. In addition, for all these diseases there has been an increase in the use of immunomodulators or immunosuppressant therapies for example in some malignancies – checkpoint inhibitors^36-38^ and CAR-T^39^; in inflammatory rheumatic diseases and IBD – biologics such as anti-Tumour Necrosis Factor antibodies and thiopurines are recommended^11,12^. Another notable finding is the relatively high numbers of patients with HLH who had co-occurring vasculitis, which may reflect a risk associated with thiopurine exposure which is often used for maintenance therapy after initial remission induction^40^. Lastly, the age at which EBV is first acquired is rising^41,42^. If this infection is first contracted or reactivates during treatment for an HLH associated comorbidity this may precipitate HLH^13^. Taken together it seems likely that some of the explanation for the increase in HLH among children over the age of 5 years and young, middle and older aged adults is due to the trends in occurrence of associated comorbidities and the changes in their treatment.

In conclusion, we have provided the first nationwide, population-based estimates of the incidence of HLH across all ages and demonstrated important temporal trends by both age and HLH-associated underlying diseases. These trends imply that some of the rise in frequency of the diagnosis of HLH may be due to either underlying diseases or their treatments and as such highlight areas in which early diagnosis or preventative measures could be developed.

## Supporting information

Supplementary Figure 1 - study flow diagram

## Data Availability

Data for this study were collected and analysed under the National Disease Registries Directions 2021, made in accordance with sections 254(1) and 254(6) of the 2012 Health and Social Care Act. Further ethical approval for this study was not required per the definition of research according to the UK Policy Framework for Health and Social Care Research. The data are available to those that have the legal basis to access it, either through the Data Access Request Service (https://digital.nhs.uk/services/data-access-request-service-dars) or partnership with National Disease Registration Service (NDRS). The protocol was approved by the joint NDRS project board reference PPF1920_027.

## Acknowledgements

We are grateful to the late Johann Visser for assistance in gaining funding for this project. This work uses data that has been provided by patients, the NHS and other health care organisations as part of routine patient care and support. The data is collated, maintained, and quality assured by the National Disease Registration Service, which are is part of NHS Digital.

## Contributions

All authors were involved in the conceptualization, acquisition of funding, drafting of the manuscript and approval of final draft for submission. PS, CJC and JW had full access to the data in the study and carried out the design of and execution of the analysis. JW had final responsibility for the decision to submit for publication.

---

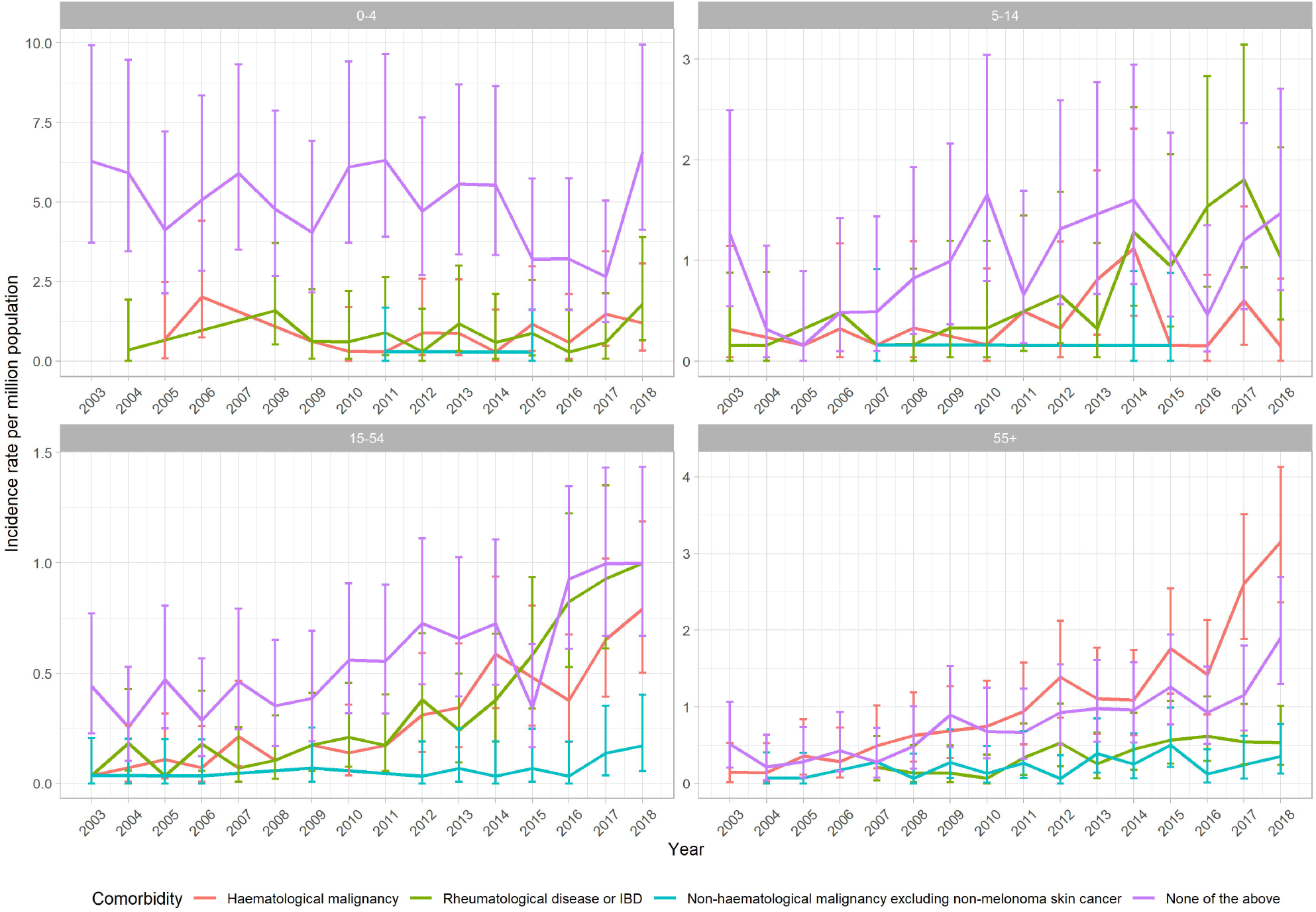

