## Supplementary figures and images for "Temporal trends in the incidence of haemophagocytic lymphohistiocytosis: a nationwide cohort study from England 2003-2018"

### Supplementary Figure 1 - study flow diagram

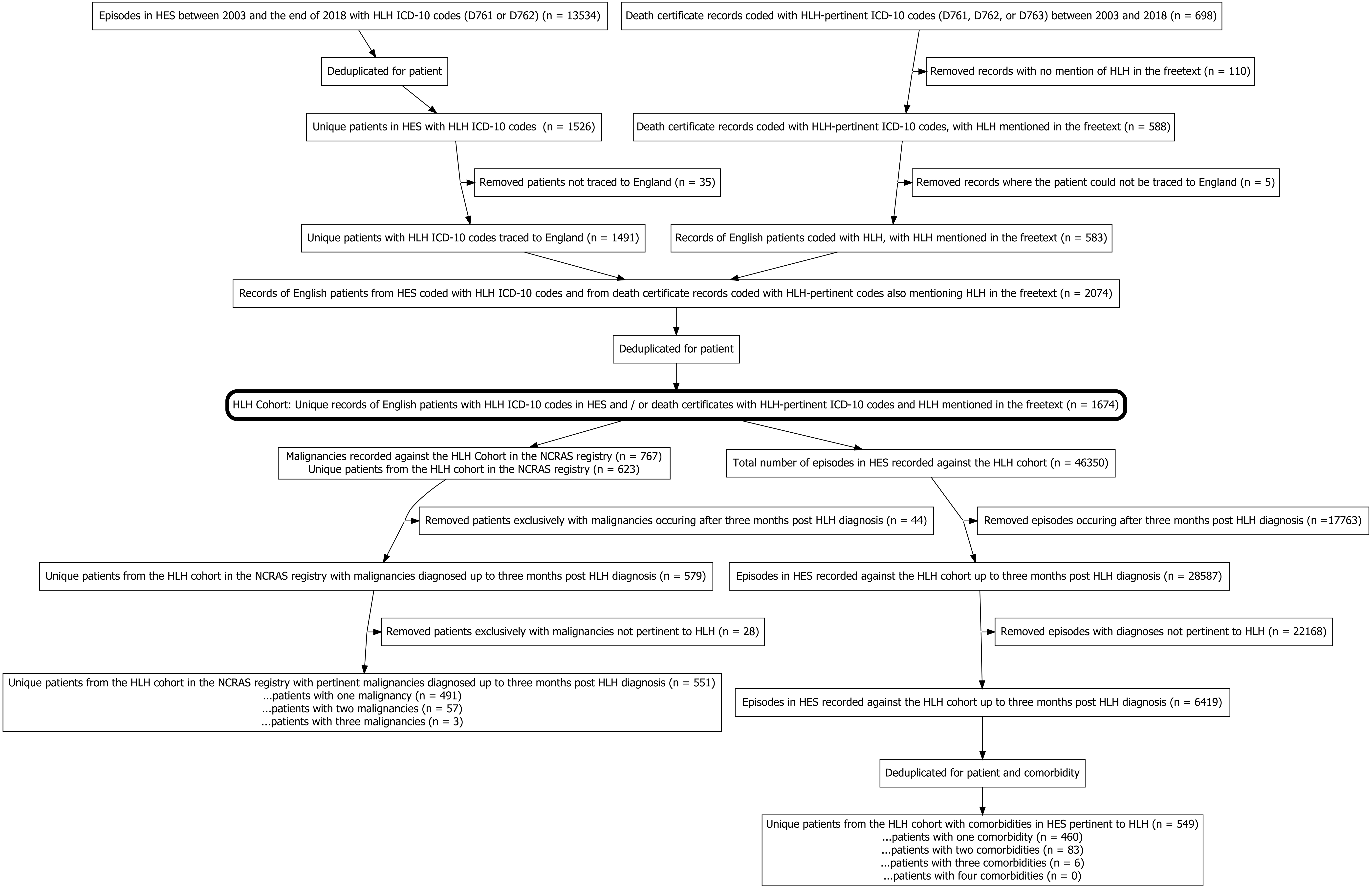
