## Supplementary Table 1 for "Temporal trends in the incidence of haemophagocytic lymphohistiocytosis: a nationwide cohort study from England 2003-2018"

Table S1. ICD 10 codes for identification and classification of associated comorbidities

| ICD-10 code | ICD-10 description | Category | Subcategory |
| --- | --- | --- | --- |
| B20 | Human immunodeficiency virus [HIV] disease resulting in infectious and parasitic diseases | Human immunodeficiency virus and Acquired Immunodeficiency Disease |  |
| B201 | HIV disease resulting in other bacterial infections | Human immunodeficiency virus and Acquired Immunodeficiency Disease |  |
| B202 | HIV disease resulting in cytomegaloviral disease | Human immunodeficiency virus and Acquired Immunodeficiency Disease |  |
| B203 | HIV disease resulting in other viral infections | Human immunodeficiency virus and Acquired Immunodeficiency Disease |  |
| B204 | HIV disease resulting in candidiasis | Human immunodeficiency virus and Acquired Immunodeficiency Disease |  |
| B205 | HIV disease resulting in other mycoses | Human immunodeficiency virus and Acquired Immunodeficiency Disease |  |
| B206 | HIV disease resulting in Pneumocystis jirovecii pneumonia | Human immunodeficiency virus and Acquired Immunodeficiency Disease |  |
| B207 | HIV disease resulting in multiple infections | Human immunodeficiency virus and Acquired Immunodeficiency Disease |  |
| B208 | HIV disease resulting in other infectious and parasitic diseases | Human immunodeficiency virus and Acquired Immunodeficiency Disease |  |
| B209 | HIV disease resulting in unspecified infectious or parasitic disease | Human immunodeficiency virus and Acquired Immunodeficiency Disease |  |
| B210 | HIV disease resulting in Kaposi sarcoma | Human immunodeficiency virus and Acquired Immunodeficiency Disease |  |
| B211 | HIV disease resulting in Burkitt lymphoma | Human immunodeficiency virus and Acquired Immunodeficiency Disease |  |
| B212 | HIV disease resulting in other types of non-Hodgkin lymphoma | Human immunodeficiency virus and Acquired Immunodeficiency Disease |  |
| B213 | HIV disease resulting in other malignant neoplasms of lymphoid, haematopoietic and related tissue | Human immunodeficiency virus and Acquired Immunodeficiency Disease |  |
| B217 | HIV disease resulting in multiple malignant neoplasms | Human immunodeficiency virus and Acquired Immunodeficiency Disease |  |
| B218 | HIV disease resulting in other malignant neoplasms | Human immunodeficiency virus and Acquired Immunodeficiency Disease |  |
| B219 | HIV disease resulting in unspecified malignant neoplasm | Human immunodeficiency virus and Acquired Immunodeficiency Disease |  |
| B220 | HIV disease resulting in encephalopathy | Human immunodeficiency virus and Acquired Immunodeficiency Disease |  |
| B221 | HIV disease resulting in lymphoid interstitial pneumonitis | Human immunodeficiency virus and Acquired Immunodeficiency Disease |  |
| B222 | HIV disease resulting in wasting syndrome | Human immunodeficiency virus and Acquired Immunodeficiency Disease |  |
| B227 | HIV disease resulting in multiple diseases classified elsewhere | Human immunodeficiency virus and Acquired Immunodeficiency Disease |  |
| B230 | Acute HIV infection syndrome | Human immunodeficiency virus and Acquired Immunodeficiency Disease |  |
| B231 | HIV disease resulting in (persistent) generalized lymphadenopathy | Human immunodeficiency virus and Acquired Immunodeficiency Disease |  |
| B232 | HIV disease resulting in haematological and immunological abnormalities, not elsewhere classified | Human immunodeficiency virus and Acquired Immunodeficiency Disease |  |
| B238 | HIV disease resulting in other specified conditions | Human immunodeficiency virus and Acquired Immunodeficiency Disease |  |
| B24 | Unspecified human immunodeficiency virus [HIV] disease | Human immunodeficiency virus and Acquired Immunodeficiency Disease |  |
| F024 | Dementia in human immunodeficiency virus [HIV] disease | Human immunodeficiency virus and Acquired Immunodeficiency Disease |  |
| O987 | Human immunodeficiency virus [HIV] disease complicating pregnancy, childbirth and the puerperium | Human immunodeficiency virus and Acquired Immunodeficiency Disease |  |
| R75 | Laboratory evidence of human immunodeficiency virus [HIV] | Human immunodeficiency virus and Acquired Immunodeficiency Disease |  |
| Z21 | Asymptomatic human immunodeficiency virus [HIV] infection status | Human immunodeficiency virus and Acquired Immunodeficiency Disease |  |
| Z717 | Human immunodeficiency virus [HIV] counselling | Human immunodeficiency virus and Acquired Immunodeficiency Disease |  |
| B200 | Human immunodeficiency virus [HIV] disease resulting in infectious and parasitic diseases | Human immunodeficiency virus and Acquired Immunodeficiency Disease |  |
| B240 | Unspecified human immunodeficiency virus [HIV] disease | Human immunodeficiency virus and Acquired Immunodeficiency Disease |  |
| R750 | Laboratory evidence of human immunodeficiency virus [HIV] | Human immunodeficiency virus and Acquired Immunodeficiency Disease |  |
| Z210 | Asymptomatic human immunodeficiency virus [HIV] infection status | Human immunodeficiency virus and Acquired Immunodeficiency Disease |  |
| B20X | Human immunodeficiency virus [HIV] disease resulting in infectious and parasitic diseases | Human immunodeficiency virus and Acquired Immunodeficiency Disease |  |
| B24X | Unspecified human immunodeficiency virus [HIV] disease | Human immunodeficiency virus and Acquired Immunodeficiency Disease |  |
| R75X | Laboratory evidence of human immunodeficiency virus [HIV] | Human immunodeficiency virus and Acquired Immunodeficiency Disease |  |
| Z21X | Asymptomatic human immunodeficiency virus [HIV] infection status | Human immunodeficiency virus and Acquired Immunodeficiency Disease |  |
| K500 | Crohn disease of small intestine | Inflammatory bowel disease |  |
| K501 | Crohn disease of large intestine | Inflammatory bowel disease |  |
| K508 | Other Crohn disease | Inflammatory bowel disease |  |
| K509 | Crohn disease, unspecified | Inflammatory bowel disease |  |
| K510 | Ulcerative (chronic) pancolitis | Inflammatory bowel disease |  |
| K512 | Ulcerative (chronic) proctitis | Inflammatory bowel disease |  |
| K513 | Ulcerative (chronic) rectosigmoiditis | Inflammatory bowel disease |  |
| K514 | Inflammatory polyps | Inflammatory bowel disease |  |
| K515 | Left sided colitis | Inflammatory bowel disease |  |
| K518 | Other ulcerative colitis | Inflammatory bowel disease |  |
| K519 | Ulcerative colitis, unspecified | Inflammatory bowel disease |  |
| K520 | Gastroenteritis and colitis due to radiation | Inflammatory bowel disease |  |
| K521 | Toxic gastroenteritis and colitis | Inflammatory bowel disease |  |
| K522 | Allergic and dietetic gastroenteritis and colitis | Inflammatory bowel disease |  |
| K523 | Indeterminate colitis | Inflammatory bowel disease |  |
| K528 | Other specified noninfective gastroenteritis and colitis | Inflammatory bowel disease |  |
| M061 | Adult-onset Still disease | Rheumatological disease | Adult-onset Still disease |
| M023 | Reiter's disease | Rheumatological disease | Inflammatory arthritis |
| M028 | Other reactive arthropathies | Rheumatological disease | Inflammatory arthritis |
| M029 | Reactive arthropathy, unspecified | Rheumatological disease | Inflammatory arthritis |
| M036 | Reactive arthropathy in other diseases classified elsewhere | Rheumatological disease | Inflammatory arthritis |
| M070 | Distal interphalangeal psoriatic arthropathy | Rheumatological disease | Inflammatory arthritis |
| M071 | Arthritis mutilans | Rheumatological disease | Inflammatory arthritis |
| M072 | Psoriatic spondylitis | Rheumatological disease | Inflammatory arthritis |
| M073 | Other psoriatic arthropathies | Rheumatological disease | Inflammatory arthritis |
| M072 | Psoriatic spondylitis | Rheumatological disease | Inflammatory arthritis |
| M45X | Ankylosing spondylitis | Rheumatological disease | Inflammatory arthritis |
| M488 | Other specified spondylopathies | Rheumatological disease | Inflammatory arthritis |
| M489 | Spondylopathy, unspecified | Rheumatological disease | Inflammatory arthritis |
| D891 | Cryoglobulinaemia | Rheumatological disease | Other connective tissue diseases |
| G724 | Inflammatory myopathy, not elsewhere classified | Rheumatological disease | Other connective tissue diseases |
| J991 | Respiratory disorders in other diffuse connective tissue disorders | Rheumatological disease | Other connective tissue diseases |
| M330 | Juvenile dermatomyositis | Rheumatological disease | Other connective tissue diseases |
| M331 | Other dermatomyositis | Rheumatological disease | Other connective tissue diseases |
| M332 | Polymyositis | Rheumatological disease | Other connective tissue diseases |
| M339 | Dermatopolymyositis, unspecified | Rheumatological disease | Other connective tissue diseases |
| M340 | Progressive systemic sclerosis | Rheumatological disease | Other connective tissue diseases |
| M341 | CR(E)ST syndrome | Rheumatological disease | Other connective tissue diseases |
| M342 | Systemic sclerosis induced by drugs and chemicals | Rheumatological disease | Other connective tissue diseases |
| M348 | Other forms of systemic sclerosis | Rheumatological disease | Other connective tissue diseases |
| M349 | Systemic sclerosis, unspecified | Rheumatological disease | Other connective tissue diseases |
| M350 | Sicca syndrome [Sjögren] | Rheumatological disease | Other connective tissue diseases |
| M351 | Other overlap syndromes | Rheumatological disease | Other connective tissue diseases |
| M354 | Diffuse (eosinophilic) fasciitis | Rheumatological disease | Other connective tissue diseases |
| M356 | Relapsing panniculitis [Weber-Christian] | Rheumatological disease | Other connective tissue diseases |
| M357 | Hypermobility syndrome | Rheumatological disease | Other connective tissue diseases |
| M358 | Other specified systemic involvement of connective tissue | Rheumatological disease | Other connective tissue diseases |
| M359 | Systemic involvement of connective tissue, unspecified | Rheumatological disease | Other connective tissue diseases |
| M368 | Systemic disorders of connective tissue in other diseases classified elsewhere | Rheumatological disease | Other connective tissue diseases |
| M608 | Other myositis | Rheumatological disease | Other connective tissue diseases |
| M609 | Myositis, unspecified | Rheumatological disease | Other connective tissue diseases |
| N082 | Glomerular disorders in blood diseases and disorders involving the immune mechanism | Rheumatological disease | Other connective tissue diseases |
| N085 | Glomerular disorders in systemic connective tissue disorders | Rheumatological disease | Other connective tissue diseases |
| N164 | Renal tubulo-interstitial disorders in systemic connective tissue disorders | Rheumatological disease | Other connective tissue diseases |
| J990 | Rheumatoid lung disease | Rheumatological disease | Rheumatoid arthritis |
| M05 | Seropositive rheumatoid arthritis | Rheumatological disease | Rheumatoid arthritis |
| M050 | Felty syndrome | Rheumatological disease | Rheumatoid arthritis |
| M050 | Seropositive rheumatoid arthritis | Rheumatological disease | Rheumatoid arthritis |
| M051 | Seropositive rheumatoid arthritis: Rheumatoid lung disease | Rheumatological disease | Rheumatoid arthritis |
| M052 | Rheumatoid vasculitis | Rheumatological disease | Rheumatoid arthritis |
| M053 | Rheumatoid arthritis with involvement of other organs and systems | Rheumatological disease | Rheumatoid arthritis |
| M058 | Other seropositive rheumatoid arthritis | Rheumatological disease | Rheumatoid arthritis |
| M059 | Seropositive rheumatoid arthritis, unspecified | Rheumatological disease | Rheumatoid arthritis |
| M05X | Seropositive rheumatoid arthritis | Rheumatological disease | Rheumatoid arthritis |
| M060 | Seronegative rheumatoid arthritis | Rheumatological disease | Rheumatoid arthritis |
| M062 | Rheumatoid bursitis | Rheumatological disease | Rheumatoid arthritis |
| M063 | Rheumatoid nodule | Rheumatological disease | Rheumatoid arthritis |
| M064 | Inflammatory polyarthropathy | Rheumatological disease | Rheumatoid arthritis |
| M068 | Other specified rheumatoid arthritis | Rheumatological disease | Rheumatoid arthritis |
| M069 | Rheumatoid arthritis, unspecified | Rheumatological disease | Rheumatoid arthritis |
| M08 | Juvenile arthritis | Rheumatological disease | Systemic juvenile idiopathic arthritis |
| M080 | Juvenile rheumatoid arthritis | Rheumatological disease | Systemic juvenile idiopathic arthritis |
| M080 | Juvenile arthritis | Rheumatological disease | Systemic juvenile idiopathic arthritis |
| M081 | Juvenile ankylosing spondylitis | Rheumatological disease | Systemic juvenile idiopathic arthritis |
| M082 | Juvenile arthritis with systemic onset | Rheumatological disease | Systemic juvenile idiopathic arthritis |
| M083 | Juvenile polyarthritis (seronegative) | Rheumatological disease | Systemic juvenile idiopathic arthritis |
| M084 | Pauciarticular juvenile arthritis | Rheumatological disease | Systemic juvenile idiopathic arthritis |
| M088 | Other juvenile arthritis | Rheumatological disease | Systemic juvenile idiopathic arthritis |
| M089 | Juvenile arthritis, unspecified | Rheumatological disease | Systemic juvenile idiopathic arthritis |
| M08X | Juvenile arthritis | Rheumatological disease | Systemic juvenile idiopathic arthritis |
| M090 | Juvenile arthritis in psoriasis | Rheumatological disease | Systemic juvenile idiopathic arthritis |
| M0980 | Juvenile arthritis in other diseases classified elsewhere | Rheumatological disease | Systemic juvenile idiopathic arthritis |
| L930 | Lupus erythematous (discoid) (NOS) | Rheumatological disease | Systemic lupus erythematosus |
| M32 | SLE | Rheumatological disease | Systemic lupus erythematosus |
| M320 | Drug-induced systemic lupus erythematosus | Rheumatological disease | Systemic lupus erythematosus |
| M321 | Systemic lupus erythematosus with organ or system involvement | Rheumatological disease | Systemic lupus erythematosus |
| M328 | Other forms of systemic lupus erythematosus | Rheumatological disease | Systemic lupus erythematosus |
| M328 | Other forms of SLE | Rheumatological disease | Systemic lupus erythematosus |
| M329 | Systemic lupus erythematosus, unspecified | Rheumatological disease | Systemic lupus erythematosus |
| M329 | SLE, unspecified | Rheumatological disease | Systemic lupus erythematosus |
| I776 | Arteritis, unspecified | Rheumatological disease | Vasculitis |
| I778 | Other specified disorders of arteries and arterioles | Rheumatological disease | Vasculitis |
| I779 | Disorder of arteries and arterioles, unspecified | Rheumatological disease | Vasculitis |
| L950 | Livedoid vasculitis | Rheumatological disease | Vasculitis |
| L958 | Other vasculitis limited to skin | Rheumatological disease | Vasculitis |
| L959 | Vasculitis limited to skin, unspecified | Rheumatological disease | Vasculitis |
| M300 | Polyarteritis nodosa | Rheumatological disease | Vasculitis |
| M301 | Polyarteritis with lung involvement [Churg-Strauss] | Rheumatological disease | Vasculitis |
| M302 | Juvenile polyarteritis | Rheumatological disease | Vasculitis |
| M303 | Mucocutaneous lymph node syndrome [Kawasaki] | Rheumatological disease | Vasculitis |
| M308 | Other conditions related to polyarteritis nodosa | Rheumatological disease | Vasculitis |
| M310 | Hypersensitivity angiitis | Rheumatological disease | Vasculitis |
| M311 | Thrombotic microangiopathy | Rheumatological disease | Vasculitis |
| M312 | Lethal midline granuloma | Rheumatological disease | Vasculitis |
| M313 | Wegener granulomatosis | Rheumatological disease | Vasculitis |
| M314 | Aortic arch syndrome [Takayasu] | Rheumatological disease | Vasculitis |
| M315 | Giant cell arteritis with polymyalgia rheumatica | Rheumatological disease | Vasculitis |
| M316 | Other giant cell arteritis | Rheumatological disease | Vasculitis |
| M317 | Microscopic polyangiitis | Rheumatological disease | Vasculitis |
| M318 | Other specified necrotizing vasculopathies | Rheumatological disease | Vasculitis |
| M319 | Necrotizing vasculopathy, unspecified | Rheumatological disease | Vasculitis |
| M352 | Behçet disease | Rheumatological disease | Vasculitis |
| M353 | Polymyalgia rheumatica | Rheumatological disease | Vasculitis |
| M941 | Relapsing polychondritis | Rheumatological disease | Vasculitis |
