## Supplementary Table 2 for "Temporal trends in the incidence of haemophagocytic lymphohistiocytosis: a nationwide cohort study from England 2003-2018"

Table S2. ICD 10 codes to identify herpes viruses

| ICD-10 code | ICD-10 description | Category |
| --- | --- | --- |
| B25 | Cytomegaloviral disease | Cytomegalovirus |
| B250 | Cytomegaloviral pneumonitis | Cytomegalovirus |
| B251 | Cytomegaloviral hepatitis | Cytomegalovirus |
| B252 | Cytomegaloviral pancreatitis | Cytomegalovirus |
| B258 | Other cytomegaloviral diseases | Cytomegalovirus |
| B259 | Cytomegaloviral disease, unspecified | Cytomegalovirus |
| B271 | Cytomegaloviral mononucleosis | Cytomegalovirus |
| J171 | Pneumonia in viral diseases classified elsewhere | Cytomegalovirus |
| K871 | Disorders of pancreas in diseases classified elsewhere | Cytomegalovirus |
| P351 | Congenital cytomegalovirus infection | Cytomegalovirus |
| B02 | Zoster [herpes zoster] | Herpes Zoster Virus |
| B020 | Zoster encephalitis | Herpes Zoster Virus |
| B021 | Zoster meningitis | Herpes Zoster Virus |
| B023 | Zoster ocular disease | Herpes Zoster Virus |
| B027 | Disseminated zoster | Herpes Zoster Virus |
| B028 | Zoster with other complications | Herpes Zoster Virus |
| B029 | Zoster without complication | Herpes Zoster Virus |
| B270 | Gammaherpesviral mononucleosis | Epstein-Barr Virus |
| B279 | Infectious mononucleosis, unspecified | Epstein-Barr Virus |
| D823 | Immunodeficiency following hereditary defective response to Epstein-Barr virus | Epstein-Barr Virus |
| B250 | Cytomegaloviral disease | Cytomegalovirus |
| B020 | Zoster [herpes zoster] | Herpes Zoster Virus |
| B25X | Cytomegaloviral disease | Cytomegalovirus |
| B02X | Zoster [herpes zoster] | Herpes Zoster Virus |
