## Supplementary Table 3 for "Temporal trends in the incidence of haemophagocytic lymphohistiocytosis: a nationwide cohort study from England 2003-2018"

Table S3. ICD 10 codes to malignancy

| ICD-10 code | ICD-10 description | Category | Hierarchical grouping |
| --- | --- | --- | --- |
| C000 | Malignant neoplasm: External upper lip | Non haematological malignancy excluding non melonoma skin cancer | Non haematological malignancy |
| C001 | Malignant neoplasm: External lower lip | Non haematological malignancy excluding non melonoma skin cancer | Non haematological malignancy |
| C002 | Malignant neoplasm: External lip, unspecified | Non haematological malignancy excluding non melonoma skin cancer | Non haematological malignancy |
| C003 | Malignant neoplasm: Upper lip, inner aspect | Non haematological malignancy excluding non melonoma skin cancer | Non haematological malignancy |
| C004 | Malignant neoplasm: Lower lip, inner aspect | Non haematological malignancy excluding non melonoma skin cancer | Non haematological malignancy |
| C005 | Malignant neoplasm: Lip, unspecified, inner aspect | Non haematological malignancy excluding non melonoma skin cancer | Non haematological malignancy |
| C006 | Malignant neoplasm: Commissure of lip | Non haematological malignancy excluding non melonoma skin cancer | Non haematological malignancy |
| C008 | Malignant neoplasm: Overlapping lesion of lip | Non haematological malignancy excluding non melonoma skin cancer | Non haematological malignancy |
| C009 | Malignant neoplasm: Lip, unspecified | Non haematological malignancy excluding non melonoma skin cancer | Non haematological malignancy |
| C01 | Malignant neoplasm of base of tongue | Non haematological malignancy excluding non melonoma skin cancer | Non haematological malignancy |
| C010 | Malignant neoplasm of base of tongue | Non haematological malignancy excluding non melonoma skin cancer | Non haematological malignancy |
| C01X | Malignant neoplasm of base of tongue | Non haematological malignancy excluding non melonoma skin cancer | Non haematological malignancy |
| C020 | Malignant neoplasm: Dorsal surface of tongue | Non haematological malignancy excluding non melonoma skin cancer | Non haematological malignancy |
| C021 | Malignant neoplasm: Border of tongue | Non haematological malignancy excluding non melonoma skin cancer | Non haematological malignancy |
| C022 | Malignant neoplasm: Ventral surface of tongue | Non haematological malignancy excluding non melonoma skin cancer | Non haematological malignancy |
| C023 | Malignant neoplasm: Anterior two-thirds of tongue, part unspecified | Non haematological malignancy excluding non melonoma skin cancer | Non haematological malignancy |
| C024 | Malignant neoplasm: Lingual tonsil | Non haematological malignancy excluding non melonoma skin cancer | Non haematological malignancy |
| C028 | Malignant neoplasm: Overlapping lesion of tongue | Non haematological malignancy excluding non melonoma skin cancer | Non haematological malignancy |
| C029 | Malignant neoplasm: Tongue, unspecified | Non haematological malignancy excluding non melonoma skin cancer | Non haematological malignancy |
| C030 | Malignant neoplasm: Upper gum | Non haematological malignancy excluding non melonoma skin cancer | Non haematological malignancy |
| C031 | Malignant neoplasm: Lower gum | Non haematological malignancy excluding non melonoma skin cancer | Non haematological malignancy |
| C039 | Malignant neoplasm: Gum, unspecified | Non haematological malignancy excluding non melonoma skin cancer | Non haematological malignancy |
| C040 | Malignant neoplasm: Anterior floor of mouth | Non haematological malignancy excluding non melonoma skin cancer | Non haematological malignancy |
| C041 | Malignant neoplasm: Lateral floor of mouth | Non haematological malignancy excluding non melonoma skin cancer | Non haematological malignancy |
| C048 | Malignant neoplasm: Overlapping lesion of floor of mouth | Non haematological malignancy excluding non melonoma skin cancer | Non haematological malignancy |
| C049 | Malignant neoplasm: Floor of mouth, unspecified | Non haematological malignancy excluding non melonoma skin cancer | Non haematological malignancy |
| C050 | Malignant neoplasm: Hard palate | Non haematological malignancy excluding non melonoma skin cancer | Non haematological malignancy |
| C051 | Malignant neoplasm: Soft palate | Non haematological malignancy excluding non melonoma skin cancer | Non haematological malignancy |
| C052 | Malignant neoplasm: Uvula | Non haematological malignancy excluding non melonoma skin cancer | Non haematological malignancy |
| C058 | Malignant neoplasm: Overlapping lesion of palate | Non haematological malignancy excluding non melonoma skin cancer | Non haematological malignancy |
| C059 | Malignant neoplasm: Palate, unspecified | Non haematological malignancy excluding non melonoma skin cancer | Non haematological malignancy |
| C060 | Malignant neoplasm: Cheek mucosa | Non haematological malignancy excluding non melonoma skin cancer | Non haematological malignancy |
| C061 | Malignant neoplasm: Vestibule of mouth | Non haematological malignancy excluding non melonoma skin cancer | Non haematological malignancy |
| C062 | Malignant neoplasm: Retromolar area | Non haematological malignancy excluding non melonoma skin cancer | Non haematological malignancy |
| C068 | Malignant neoplasm: Overlapping lesion of other and unspecified parts of mouth | Non haematological malignancy excluding non melonoma skin cancer | Non haematological malignancy |
| C069 | Malignant neoplasm: Mouth, unspecified | Non haematological malignancy excluding non melonoma skin cancer | Non haematological malignancy |
| C07 | Malignant neoplasm of parotid gland | Non haematological malignancy excluding non melonoma skin cancer | Non haematological malignancy |
| C070 | Malignant neoplasm of parotid gland | Non haematological malignancy excluding non melonoma skin cancer | Non haematological malignancy |
| C07X | Malignant neoplasm of parotid gland | Non haematological malignancy excluding non melonoma skin cancer | Non haematological malignancy |
| C080 | Malignant neoplasm: Submandibular gland | Non haematological malignancy excluding non melonoma skin cancer | Non haematological malignancy |
| C081 | Malignant neoplasm: Sublingual gland | Non haematological malignancy excluding non melonoma skin cancer | Non haematological malignancy |
| C088 | Malignant neoplasm: Overlapping lesion of major salivary glands | Non haematological malignancy excluding non melonoma skin cancer | Non haematological malignancy |
| C089 | Malignant neoplasm: Major salivary gland, unspecified | Non haematological malignancy excluding non melonoma skin cancer | Non haematological malignancy |
| C090 | Malignant neoplasm: Tonsillar fossa | Non haematological malignancy excluding non melonoma skin cancer | Non haematological malignancy |
| C091 | Malignant neoplasm: Tonsillar pillar (anterior)(posterior) | Non haematological malignancy excluding non melonoma skin cancer | Non haematological malignancy |
| C098 | Malignant neoplasm: Overlapping lesion of tonsil | Non haematological malignancy excluding non melonoma skin cancer | Non haematological malignancy |
| C099 | Malignant neoplasm: Tonsil, unspecified | Non haematological malignancy excluding non melonoma skin cancer | Non haematological malignancy |
| C100 | Malignant neoplasm: Vallecula | Non haematological malignancy excluding non melonoma skin cancer | Non haematological malignancy |
| C101 | Malignant neoplasm: Anterior surface of epiglottis | Non haematological malignancy excluding non melonoma skin cancer | Non haematological malignancy |
| C102 | Malignant neoplasm: Lateral wall of oropharynx | Non haematological malignancy excluding non melonoma skin cancer | Non haematological malignancy |
| C103 | Malignant neoplasm: Posterior wall of oropharynx | Non haematological malignancy excluding non melonoma skin cancer | Non haematological malignancy |
| C104 | Malignant neoplasm: Branchial cleft | Non haematological malignancy excluding non melonoma skin cancer | Non haematological malignancy |
| C108 | Malignant neoplasm: Overlapping lesion of oropharynx | Non haematological malignancy excluding non melonoma skin cancer | Non haematological malignancy |
| C109 | Malignant neoplasm: Oropharynx, unspecified | Non haematological malignancy excluding non melonoma skin cancer | Non haematological malignancy |
| C110 | Malignant neoplasm: Superior wall of nasopharynx | Non haematological malignancy excluding non melonoma skin cancer | Non haematological malignancy |
| C111 | Malignant neoplasm: Posterior wall of nasopharynx | Non haematological malignancy excluding non melonoma skin cancer | Non haematological malignancy |
| C112 | Malignant neoplasm: Lateral wall of nasopharynx | Non haematological malignancy excluding non melonoma skin cancer | Non haematological malignancy |
| C113 | Malignant neoplasm: Anterior wall of nasopharynx | Non haematological malignancy excluding non melonoma skin cancer | Non haematological malignancy |
| C118 | Malignant neoplasm: Overlapping lesion of nasopharynx | Non haematological malignancy excluding non melonoma skin cancer | Non haematological malignancy |
| C119 | Malignant neoplasm: Nasopharynx, unspecified | Non haematological malignancy excluding non melonoma skin cancer | Non haematological malignancy |
| C12 | Malignant neoplasm of piriform sinus | Non haematological malignancy excluding non melonoma skin cancer | Non haematological malignancy |
| C120 | Malignant neoplasm of pyriform sinus | Non haematological malignancy excluding non melonoma skin cancer | Non haematological malignancy |
| C12X | Malignant neoplasm of piriform sinus | Non haematological malignancy excluding non melonoma skin cancer | Non haematological malignancy |
| C130 | Malignant neoplasm: Postcricoid region | Non haematological malignancy excluding non melonoma skin cancer | Non haematological malignancy |
| C131 | Malignant neoplasm: Aryepiglottic fold, hypopharyngeal aspect | Non haematological malignancy excluding non melonoma skin cancer | Non haematological malignancy |
| C132 | Malignant neoplasm: Posterior wall of hypopharynx | Non haematological malignancy excluding non melonoma skin cancer | Non haematological malignancy |
| C138 | Malignant neoplasm: Overlapping lesion of hypopharynx | Non haematological malignancy excluding non melonoma skin cancer | Non haematological malignancy |
| C139 | Malignant neoplasm: Hypopharynx, unspecified | Non haematological malignancy excluding non melonoma skin cancer | Non haematological malignancy |
| C140 | Malignant neoplasm: Pharynx, unspecified | Non haematological malignancy excluding non melonoma skin cancer | Non haematological malignancy |
| C142 | Malignant neoplasm: Waldeyer ring | Non haematological malignancy excluding non melonoma skin cancer | Non haematological malignancy |
| C148 | Malignant neoplasm: Overlapping lesion of lip, oral cavity and pharynx | Non haematological malignancy excluding non melonoma skin cancer | Non haematological malignancy |
| C150 | Malignant neoplasm: Cervical part of oesophagus | Non haematological malignancy excluding non melonoma skin cancer | Non haematological malignancy |
| C151 | Malignant neoplasm: Thoracic part of oesophagus | Non haematological malignancy excluding non melonoma skin cancer | Non haematological malignancy |
| C152 | Malignant neoplasm: Abdominal part of oesophagus | Non haematological malignancy excluding non melonoma skin cancer | Non haematological malignancy |
| C153 | Malignant neoplasm: Upper third of oesophagus | Non haematological malignancy excluding non melonoma skin cancer | Non haematological malignancy |
| C154 | Malignant neoplasm: Middle third of oesophagus | Non haematological malignancy excluding non melonoma skin cancer | Non haematological malignancy |
| C155 | Malignant neoplasm: Lower third of oesophagus | Non haematological malignancy excluding non melonoma skin cancer | Non haematological malignancy |
| C158 | Malignant neoplasm: Overlapping lesion of oesophagus | Non haematological malignancy excluding non melonoma skin cancer | Non haematological malignancy |
| C159 | Malignant neoplasm: Oesophagus, unspecified | Non haematological malignancy excluding non melonoma skin cancer | Non haematological malignancy |
| C160 | Malignant neoplasm: Cardia | Non haematological malignancy excluding non melonoma skin cancer | Non haematological malignancy |
| C161 | Malignant neoplasm: Fundus of stomach | Non haematological malignancy excluding non melonoma skin cancer | Non haematological malignancy |
| C162 | Malignant neoplasm: Body of stomach | Non haematological malignancy excluding non melonoma skin cancer | Non haematological malignancy |
| C163 | Malignant neoplasm: Pyloric antrum | Non haematological malignancy excluding non melonoma skin cancer | Non haematological malignancy |
| C164 | Malignant neoplasm: Pylorus | Non haematological malignancy excluding non melonoma skin cancer | Non haematological malignancy |
| C165 | Malignant neoplasm: Lesser curvature of stomach, unspecified | Non haematological malignancy excluding non melonoma skin cancer | Non haematological malignancy |
| C166 | Malignant neoplasm: Greater curvature of stomach, unspecified | Non haematological malignancy excluding non melonoma skin cancer | Non haematological malignancy |
| C168 | Malignant neoplasm: Overlapping lesion of stomach | Non haematological malignancy excluding non melonoma skin cancer | Non haematological malignancy |
| C169 | Malignant neoplasm: Stomach, unspecified | Non haematological malignancy excluding non melonoma skin cancer | Non haematological malignancy |
| C170 | Malignant neoplasm: Duodenum | Non haematological malignancy excluding non melonoma skin cancer | Non haematological malignancy |
| C171 | Malignant neoplasm: Jejunum | Non haematological malignancy excluding non melonoma skin cancer | Non haematological malignancy |
| C172 | Malignant neoplasm: Ileum | Non haematological malignancy excluding non melonoma skin cancer | Non haematological malignancy |
| C173 | Malignant neoplasm: Meckel diverticulum | Non haematological malignancy excluding non melonoma skin cancer | Non haematological malignancy |
| C178 | Malignant neoplasm: Overlapping lesion of small intestine | Non haematological malignancy excluding non melonoma skin cancer | Non haematological malignancy |
| C179 | Malignant neoplasm: Small intestine, unspecified | Non haematological malignancy excluding non melonoma skin cancer | Non haematological malignancy |
| C180 | Malignant neoplasm: Caecum | Non haematological malignancy excluding non melonoma skin cancer | Non haematological malignancy |
| C181 | Malignant neoplasm: Appendix | Non haematological malignancy excluding non melonoma skin cancer | Non haematological malignancy |
| C182 | Malignant neoplasm: Ascending colon | Non haematological malignancy excluding non melonoma skin cancer | Non haematological malignancy |
| C183 | Malignant neoplasm: Hepatic flexure | Non haematological malignancy excluding non melonoma skin cancer | Non haematological malignancy |
| C184 | Malignant neoplasm: Transverse colon | Non haematological malignancy excluding non melonoma skin cancer | Non haematological malignancy |
| C185 | Malignant neoplasm: Splenic flexure | Non haematological malignancy excluding non melonoma skin cancer | Non haematological malignancy |
| C186 | Malignant neoplasm: Descending colon | Non haematological malignancy excluding non melonoma skin cancer | Non haematological malignancy |
| C187 | Malignant neoplasm: Sigmoid colon | Non haematological malignancy excluding non melonoma skin cancer | Non haematological malignancy |
| C188 | Malignant neoplasm: Overlapping lesion of colon | Non haematological malignancy excluding non melonoma skin cancer | Non haematological malignancy |
| C189 | Malignant neoplasm: Colon, unspecified | Non haematological malignancy excluding non melonoma skin cancer | Non haematological malignancy |
| C19 | Malignant neoplasm of rectosigmoid junction | Non haematological malignancy excluding non melonoma skin cancer | Non haematological malignancy |
| C190 | Malignant neoplasm of rectosigmoid junction | Non haematological malignancy excluding non melonoma skin cancer | Non haematological malignancy |
| C19X | Malignant neoplasm of rectosigmoid junction | Non haematological malignancy excluding non melonoma skin cancer | Non haematological malignancy |
| C20 | Malignant neoplasm of rectum | Non haematological malignancy excluding non melonoma skin cancer | Non haematological malignancy |
| C200 | Malignant neoplasm of rectum | Non haematological malignancy excluding non melonoma skin cancer | Non haematological malignancy |
| C20X | Malignant neoplasm of rectum | Non haematological malignancy excluding non melonoma skin cancer | Non haematological malignancy |
| C210 | Malignant neoplasm: Anus, unspecified | Non haematological malignancy excluding non melonoma skin cancer | Non haematological malignancy |
| C211 | Malignant neoplasm: Anal canal | Non haematological malignancy excluding non melonoma skin cancer | Non haematological malignancy |
| C212 | Malignant neoplasm: Cloacogenic zone | Non haematological malignancy excluding non melonoma skin cancer | Non haematological malignancy |
| C218 | Malignant neoplasm: Overlapping lesion of rectum, anus and anal canal | Non haematological malignancy excluding non melonoma skin cancer | Non haematological malignancy |
| C220 | Malignant neoplasm: Liver cell carcinoma | Non haematological malignancy excluding non melonoma skin cancer | Non haematological malignancy |
| C221 | Malignant neoplasm: Intrahepatic bile duct carcinoma | Non haematological malignancy excluding non melonoma skin cancer | Non haematological malignancy |
| C222 | Malignant neoplasm: Hepatoblastoma | Non haematological malignancy excluding non melonoma skin cancer | Non haematological malignancy |
| C223 | Malignant neoplasm: Angiosarcoma of liver | Non haematological malignancy excluding non melonoma skin cancer | Non haematological malignancy |
| C224 | Malignant neoplasm: Other sarcomas of liver | Non haematological malignancy excluding non melonoma skin cancer | Non haematological malignancy |
| C227 | Malignant neoplasm: Other specified carcinomas of liver | Non haematological malignancy excluding non melonoma skin cancer | Non haematological malignancy |
| C229 | Malignant neoplasm: Liver, unspecified | Non haematological malignancy excluding non melonoma skin cancer | Non haematological malignancy |
| C23 | Malignant neoplasm of gallbladder | Non haematological malignancy excluding non melonoma skin cancer | Non haematological malignancy |
| C230 | Malignant neoplasm of gallbladder | Non haematological malignancy excluding non melonoma skin cancer | Non haematological malignancy |
| C23X | Malignant neoplasm of gallbladder | Non haematological malignancy excluding non melonoma skin cancer | Non haematological malignancy |
| C240 | Malignant neoplasm: Extrahepatic bile duct | Non haematological malignancy excluding non melonoma skin cancer | Non haematological malignancy |
| C241 | Malignant neoplasm: Ampulla of Vater | Non haematological malignancy excluding non melonoma skin cancer | Non haematological malignancy |
| C248 | Malignant neoplasm: Overlapping lesion of biliary tract | Non haematological malignancy excluding non melonoma skin cancer | Non haematological malignancy |
| C249 | Malignant neoplasm: Biliary tract, unspecified | Non haematological malignancy excluding non melonoma skin cancer | Non haematological malignancy |
| C250 | Malignant neoplasm: Head of pancreas | Non haematological malignancy excluding non melonoma skin cancer | Non haematological malignancy |
| C251 | Malignant neoplasm: Body of pancreas | Non haematological malignancy excluding non melonoma skin cancer | Non haematological malignancy |
| C252 | Malignant neoplasm: Tail of pancreas | Non haematological malignancy excluding non melonoma skin cancer | Non haematological malignancy |
| C253 | Malignant neoplasm: Pancreatic duct | Non haematological malignancy excluding non melonoma skin cancer | Non haematological malignancy |
| C254 | Malignant neoplasm: Endocrine pancreas | Non haematological malignancy excluding non melonoma skin cancer | Non haematological malignancy |
| C257 | Malignant neoplasm: Other parts of pancreas | Non haematological malignancy excluding non melonoma skin cancer | Non haematological malignancy |
| C258 | Malignant neoplasm: Overlapping lesion of pancreas | Non haematological malignancy excluding non melonoma skin cancer | Non haematological malignancy |
| C259 | Malignant neoplasm: Pancreas, unspecified | Non haematological malignancy excluding non melonoma skin cancer | Non haematological malignancy |
| C260 | Malignant neoplasm: Intestinal tract, part unspecified | Non haematological malignancy excluding non melonoma skin cancer | Non haematological malignancy |
| C261 | Malignant neoplasm: Spleen | Non haematological malignancy excluding non melonoma skin cancer | Non haematological malignancy |
| C268 | Malignant neoplasm: Overlapping lesion of digestive system | Non haematological malignancy excluding non melonoma skin cancer | Non haematological malignancy |
| C269 | Malignant neoplasm: Ill-defined sites within the digestive system | Non haematological malignancy excluding non melonoma skin cancer | Non haematological malignancy |
| C300 | Malignant neoplasm: Nasal cavity | Non haematological malignancy excluding non melonoma skin cancer | Non haematological malignancy |
| C301 | Malignant neoplasm: Middle ear | Non haematological malignancy excluding non melonoma skin cancer | Non haematological malignancy |
| C310 | Malignant neoplasm: Maxillary sinus | Non haematological malignancy excluding non melonoma skin cancer | Non haematological malignancy |
| C311 | Malignant neoplasm: Ethmoidal sinus | Non haematological malignancy excluding non melonoma skin cancer | Non haematological malignancy |
| C312 | Malignant neoplasm: Frontal sinus | Non haematological malignancy excluding non melonoma skin cancer | Non haematological malignancy |
| C313 | Malignant neoplasm: Sphenoidal sinus | Non haematological malignancy excluding non melonoma skin cancer | Non haematological malignancy |
| C318 | Malignant neoplasm: Overlapping lesion of accessory sinuses | Non haematological malignancy excluding non melonoma skin cancer | Non haematological malignancy |
| C319 | Malignant neoplasm: Accessory sinus, unspecified | Non haematological malignancy excluding non melonoma skin cancer | Non haematological malignancy |
| C320 | Malignant neoplasm: Glottis | Non haematological malignancy excluding non melonoma skin cancer | Non haematological malignancy |
| C321 | Malignant neoplasm: Supraglottis | Non haematological malignancy excluding non melonoma skin cancer | Non haematological malignancy |
| C322 | Malignant neoplasm: Subglottis | Non haematological malignancy excluding non melonoma skin cancer | Non haematological malignancy |
| C323 | Malignant neoplasm: Laryngeal cartilage | Non haematological malignancy excluding non melonoma skin cancer | Non haematological malignancy |
| C328 | Malignant neoplasm: Overlapping lesion of larynx | Non haematological malignancy excluding non melonoma skin cancer | Non haematological malignancy |
| C329 | Malignant neoplasm: Larynx, unspecified | Non haematological malignancy excluding non melonoma skin cancer | Non haematological malignancy |
| C33 | Malignant neoplasm of trachea | Non haematological malignancy excluding non melonoma skin cancer | Non haematological malignancy |
| C330 | Malignant neoplasm of trachea | Non haematological malignancy excluding non melonoma skin cancer | Non haematological malignancy |
| C33X | Malignant neoplasm of trachea | Non haematological malignancy excluding non melonoma skin cancer | Non haematological malignancy |
| C340 | Malignant neoplasm: Main bronchus | Non haematological malignancy excluding non melonoma skin cancer | Non haematological malignancy |
| C341 | Malignant neoplasm: Upper lobe, bronchus or lung | Non haematological malignancy excluding non melonoma skin cancer | Non haematological malignancy |
| C342 | Malignant neoplasm: Middle lobe, bronchus or lung | Non haematological malignancy excluding non melonoma skin cancer | Non haematological malignancy |
| C343 | Malignant neoplasm: Lower lobe, bronchus or lung | Non haematological malignancy excluding non melonoma skin cancer | Non haematological malignancy |
| C348 | Malignant neoplasm: Overlapping lesion of bronchus and lung | Non haematological malignancy excluding non melonoma skin cancer | Non haematological malignancy |
| C349 | Malignant neoplasm: Bronchus or lung, unspecified | Non haematological malignancy excluding non melonoma skin cancer | Non haematological malignancy |
| C37 | Malignant neoplasm of thymus | Non haematological malignancy excluding non melonoma skin cancer | Non haematological malignancy |
| C370 | Malignant neoplasm of thymus | Non haematological malignancy excluding non melonoma skin cancer | Non haematological malignancy |
| C37X | Malignant neoplasm of thymus | Non haematological malignancy excluding non melonoma skin cancer | Non haematological malignancy |
| C380 | Malignant neoplasm: Heart | Non haematological malignancy excluding non melonoma skin cancer | Non haematological malignancy |
| C381 | Malignant neoplasm: Anterior mediastinum | Non haematological malignancy excluding non melonoma skin cancer | Non haematological malignancy |
| C382 | Malignant neoplasm: Posterior mediastinum | Non haematological malignancy excluding non melonoma skin cancer | Non haematological malignancy |
| C383 | Malignant neoplasm: Mediastinum, part unspecified | Non haematological malignancy excluding non melonoma skin cancer | Non haematological malignancy |
| C384 | Malignant neoplasm: Pleura | Non haematological malignancy excluding non melonoma skin cancer | Non haematological malignancy |
| C388 | Malignant neoplasm: Overlapping lesion of heart, mediastinum and pleura | Non haematological malignancy excluding non melonoma skin cancer | Non haematological malignancy |
| C390 | Malignant neoplasm: Upper respiratory tract, part unspecified | Non haematological malignancy excluding non melonoma skin cancer | Non haematological malignancy |
| C398 | Malignant neoplasm: Overlapping lesion of respiratory and intrathoracic organs | Non haematological malignancy excluding non melonoma skin cancer | Non haematological malignancy |
| C399 | Malignant neoplasm: Ill-defined sites within the respiratory system | Non haematological malignancy excluding non melonoma skin cancer | Non haematological malignancy |
| C400 | Malignant neoplasm: Scapula and long bones of upper limb | Non haematological malignancy excluding non melonoma skin cancer | Non haematological malignancy |
| C401 | Malignant neoplasm: Short bones of upper limb | Non haematological malignancy excluding non melonoma skin cancer | Non haematological malignancy |
| C402 | Malignant neoplasm: Long bones of lower limb | Non haematological malignancy excluding non melonoma skin cancer | Non haematological malignancy |
| C403 | Malignant neoplasm: Short bones of lower limb | Non haematological malignancy excluding non melonoma skin cancer | Non haematological malignancy |
| C408 | Malignant neoplasm: Overlapping lesion of bone and articular cartilage of limbs | Non haematological malignancy excluding non melonoma skin cancer | Non haematological malignancy |
| C409 | Malignant neoplasm: Bone and articular cartilage of limb, unspecified | Non haematological malignancy excluding non melonoma skin cancer | Non haematological malignancy |
| C410 | Malignant neoplasm: Bones of skull and face | Non haematological malignancy excluding non melonoma skin cancer | Non haematological malignancy |
| C411 | Malignant neoplasm: Mandible | Non haematological malignancy excluding non melonoma skin cancer | Non haematological malignancy |
| C412 | Malignant neoplasm: Vertebral column | Non haematological malignancy excluding non melonoma skin cancer | Non haematological malignancy |
| C413 | Malignant neoplasm: Ribs, sternum and clavicle | Non haematological malignancy excluding non melonoma skin cancer | Non haematological malignancy |
| C414 | Malignant neoplasm: Pelvic bones, sacrum and coccyx | Non haematological malignancy excluding non melonoma skin cancer | Non haematological malignancy |
| C418 | Malignant neoplasm: Overlapping lesion of bone and articular cartilage | Non haematological malignancy excluding non melonoma skin cancer | Non haematological malignancy |
| C419 | Malignant neoplasm: Bone and articular cartilage, unspecified | Other haematological histiocytic / Myelodysplastic / Malignancy / Unspecified | Haematological malignancy |
| C430 | Malignant neoplasm: Malignant melanoma of lip | Non haematological malignancy excluding non melonoma skin cancer | Non haematological malignancy |
| C431 | Malignant neoplasm: Malignant melanoma of eyelid, including canthus | Non haematological malignancy excluding non melonoma skin cancer | Non haematological malignancy |
| C432 | Malignant neoplasm: Malignant melanoma of ear and external auricular canal | Non haematological malignancy excluding non melonoma skin cancer | Non haematological malignancy |
| C433 | Malignant neoplasm: Malignant melanoma of other and unspecified parts of face | Non haematological malignancy excluding non melonoma skin cancer | Non haematological malignancy |
| C434 | Malignant neoplasm: Malignant melanoma of scalp and neck | Non haematological malignancy excluding non melonoma skin cancer | Non haematological malignancy |
| C435 | Malignant neoplasm: Malignant melanoma of trunk | Non haematological malignancy excluding non melonoma skin cancer | Non haematological malignancy |
| C436 | Malignant neoplasm: Malignant melanoma of upper limb, including shoulder | Non haematological malignancy excluding non melonoma skin cancer | Non haematological malignancy |
| C437 | Malignant neoplasm: Malignant melanoma of lower limb, including hip | Non haematological malignancy excluding non melonoma skin cancer | Non haematological malignancy |
| C438 | Malignant neoplasm: Overlapping malignant melanoma of skin | Non haematological malignancy excluding non melonoma skin cancer | Non haematological malignancy |
| C439 | Malignant neoplasm: Malignant melanoma of skin, unspecified | Non haematological malignancy excluding non melonoma skin cancer | Non haematological malignancy |
| C444 | Malignant neoplasm: Skin of scalp and neck | Non haematological malignancy excluding non melonoma skin cancer | Non haematological malignancy |
| C446 | Malignant neoplasm: Skin of upper limb, including shoulder | Other haematological histiocytic / Myelodysplastic / Malignancy / Unspecified | Haematological malignancy |
| C449 | Malignant neoplasm: Malignant neoplasm of skin, unspecified | Other haematological histiocytic / Myelodysplastic / Malignancy / Unspecified | Haematological malignancy |
| C450 | Mesothelioma of pleura | Non haematological malignancy excluding non melonoma skin cancer | Non haematological malignancy |
| C451 | Mesothelioma of peritoneum | Non haematological malignancy excluding non melonoma skin cancer | Non haematological malignancy |
| C452 | Mesothelioma of pericardium | Non haematological malignancy excluding non melonoma skin cancer | Non haematological malignancy |
| C457 | Mesothelioma of other sites | Non haematological malignancy excluding non melonoma skin cancer | Non haematological malignancy |
| C459 | Mesothelioma, unspecified | Non haematological malignancy excluding non melonoma skin cancer | Non haematological malignancy |
| C460 | Kaposi sarcoma of skin | Non haematological malignancy excluding non melonoma skin cancer | Non haematological malignancy |
| C461 | Kaposi sarcoma of soft tissue | Non haematological malignancy excluding non melonoma skin cancer | Non haematological malignancy |
| C462 | Kaposi sarcoma of palate | Non haematological malignancy excluding non melonoma skin cancer | Non haematological malignancy |
| C463 | Kaposi sarcoma of lymph nodes | Non haematological malignancy excluding non melonoma skin cancer | Non haematological malignancy |
| C467 | Kaposi sarcoma of other sites | Non haematological malignancy excluding non melonoma skin cancer | Non haematological malignancy |
| C468 | Kaposi sarcoma of multiple organs | Non haematological malignancy excluding non melonoma skin cancer | Non haematological malignancy |
| C469 | Kaposi sarcoma, unspecified | Non haematological malignancy excluding non melonoma skin cancer | Non haematological malignancy |
| C470 | Malignant neoplasm: Peripheral nerves of head, face and neck | Non haematological malignancy excluding non melonoma skin cancer | Non haematological malignancy |
| C471 | Malignant neoplasm: Peripheral nerves of upper limb, including shoulder | Non haematological malignancy excluding non melonoma skin cancer | Non haematological malignancy |
| C472 | Malignant neoplasm: Peripheral nerves of lower limb, including hip | Non haematological malignancy excluding non melonoma skin cancer | Non haematological malignancy |
| C473 | Malignant neoplasm: Peripheral nerves of thorax | Non haematological malignancy excluding non melonoma skin cancer | Non haematological malignancy |
| C474 | Malignant neoplasm: Peripheral nerves of abdomen | Non haematological malignancy excluding non melonoma skin cancer | Non haematological malignancy |
| C475 | Malignant neoplasm: Peripheral nerves of pelvis | Non haematological malignancy excluding non melonoma skin cancer | Non haematological malignancy |
| C476 | Malignant neoplasm: Peripheral nerves of trunk, unspecified | Non haematological malignancy excluding non melonoma skin cancer | Non haematological malignancy |
| C478 | Malignant neoplasm: Overlapping lesion of peripheral nerves and autonomic nervous system | Non haematological malignancy excluding non melonoma skin cancer | Non haematological malignancy |
| C479 | Malignant neoplasm: Peripheral nerves and autonomic nervous system, unspecified | Non haematological malignancy excluding non melonoma skin cancer | Non haematological malignancy |
| C480 | Malignant neoplasm: Retroperitoneum | Non haematological malignancy excluding non melonoma skin cancer | Non haematological malignancy |
| C481 | Malignant neoplasm: Specified parts of peritoneum | Non haematological malignancy excluding non melonoma skin cancer | Non haematological malignancy |
| C482 | Malignant neoplasm: Peritoneum, unspecified | Non haematological malignancy excluding non melonoma skin cancer | Non haematological malignancy |
| C488 | Malignant neoplasm: Overlapping lesion of retroperitoneum and peritoneum | Non haematological malignancy excluding non melonoma skin cancer | Non haematological malignancy |
| C490 | Malignant neoplasm: Connective and soft tissue of head, face and neck | Non haematological malignancy excluding non melonoma skin cancer | Non haematological malignancy |
| C491 | Malignant neoplasm: Connective and soft tissue of upper limb, including shoulder | Non haematological malignancy excluding non melonoma skin cancer | Non haematological malignancy |
| C492 | Malignant neoplasm: Connective and soft tissue of lower limb, including hip | Non haematological malignancy excluding non melonoma skin cancer | Non haematological malignancy |
| C493 | Malignant neoplasm: Connective and soft tissue of thorax | Non haematological malignancy excluding non melonoma skin cancer | Non haematological malignancy |
| C494 | Malignant neoplasm: Connective and soft tissue of abdomen | Non haematological malignancy excluding non melonoma skin cancer | Non haematological malignancy |
| C495 | Malignant neoplasm: Connective and soft tissue of pelvis | Non haematological malignancy excluding non melonoma skin cancer | Non haematological malignancy |
| C496 | Malignant neoplasm: Connective and soft tissue of trunk, unspecified | Non haematological malignancy excluding non melonoma skin cancer | Non haematological malignancy |
| C498 | Malignant neoplasm: Overlapping lesion of connective and soft tissue | Non haematological malignancy excluding non melonoma skin cancer | Non haematological malignancy |
| C499 | Malignant neoplasm: Connective and soft tissue, unspecified | Non haematological malignancy excluding non melonoma skin cancer | Non haematological malignancy |
| C500 | Malignant neoplasm: Nipple and areola | Non haematological malignancy excluding non melonoma skin cancer | Non haematological malignancy |
| C501 | Malignant neoplasm: Central portion of breast | Non haematological malignancy excluding non melonoma skin cancer | Non haematological malignancy |
| C502 | Malignant neoplasm: Upper-inner quadrant of breast | Non haematological malignancy excluding non melonoma skin cancer | Non haematological malignancy |
| C503 | Malignant neoplasm: Lower-inner quadrant of breast | Non haematological malignancy excluding non melonoma skin cancer | Non haematological malignancy |
| C504 | Malignant neoplasm: Upper-outer quadrant of breast | Non haematological malignancy excluding non melonoma skin cancer | Non haematological malignancy |
| C505 | Malignant neoplasm: Lower-outer quadrant of breast | Non haematological malignancy excluding non melonoma skin cancer | Non haematological malignancy |
| C506 | Malignant neoplasm: Axillary tail of breast | Non haematological malignancy excluding non melonoma skin cancer | Non haematological malignancy |
| C508 | Malignant neoplasm: Overlapping lesion of breast | Non haematological malignancy excluding non melonoma skin cancer | Non haematological malignancy |
| C509 | Malignant neoplasm: Breast, unspecified | Non haematological malignancy excluding non melonoma skin cancer | Non haematological malignancy |
| C510 | Malignant neoplasm: Labium majus | Non haematological malignancy excluding non melonoma skin cancer | Non haematological malignancy |
| C511 | Malignant neoplasm: Labium minus | Non haematological malignancy excluding non melonoma skin cancer | Non haematological malignancy |
| C512 | Malignant neoplasm: Clitoris | Non haematological malignancy excluding non melonoma skin cancer | Non haematological malignancy |
| C518 | Malignant neoplasm: Overlapping lesion of vulva | Non haematological malignancy excluding non melonoma skin cancer | Non haematological malignancy |
| C519 | Malignant neoplasm: Vulva, unspecified | Non haematological malignancy excluding non melonoma skin cancer | Non haematological malignancy |
| C52 | Malignant neoplasm of vagina | Non haematological malignancy excluding non melonoma skin cancer | Non haematological malignancy |
| C520 | Malignant neoplasm of vagina | Non haematological malignancy excluding non melonoma skin cancer | Non haematological malignancy |
| C52X | Malignant neoplasm of vagina | Non haematological malignancy excluding non melonoma skin cancer | Non haematological malignancy |
| C530 | Malignant neoplasm: Endocervix | Non haematological malignancy excluding non melonoma skin cancer | Non haematological malignancy |
| C531 | Malignant neoplasm: Exocervix | Non haematological malignancy excluding non melonoma skin cancer | Non haematological malignancy |
| C538 | Malignant neoplasm: Overlapping lesion of cervix uteri | Non haematological malignancy excluding non melonoma skin cancer | Non haematological malignancy |
| C539 | Malignant neoplasm: Cervix uteri, unspecified | Non haematological malignancy excluding non melonoma skin cancer | Non haematological malignancy |
| C540 | Malignant neoplasm: Isthmus uteri | Non haematological malignancy excluding non melonoma skin cancer | Non haematological malignancy |
| C541 | Malignant neoplasm: Endometrium | Non haematological malignancy excluding non melonoma skin cancer | Non haematological malignancy |
| C542 | Malignant neoplasm: Myometrium | Non haematological malignancy excluding non melonoma skin cancer | Non haematological malignancy |
| C543 | Malignant neoplasm: Fundus uteri | Non haematological malignancy excluding non melonoma skin cancer | Non haematological malignancy |
| C548 | Malignant neoplasm: Overlapping lesion of corpus uteri | Non haematological malignancy excluding non melonoma skin cancer | Non haematological malignancy |
| C549 | Malignant neoplasm: Corpus uteri, unspecified | Non haematological malignancy excluding non melonoma skin cancer | Non haematological malignancy |
| C55 | Malignant neoplasm of uterus, part unspecified | Non haematological malignancy excluding non melonoma skin cancer | Non haematological malignancy |
| C550 | Malignant neoplasm of uterus, part unspecified | Non haematological malignancy excluding non melonoma skin cancer | Non haematological malignancy |
| C55X | Malignant neoplasm of uterus, part unspecified | Non haematological malignancy excluding non melonoma skin cancer | Non haematological malignancy |
| C56 | Malignant neoplasm of ovary | Non haematological malignancy excluding non melonoma skin cancer | Non haematological malignancy |
| C560 | Malignant neoplasm of ovary | Non haematological malignancy excluding non melonoma skin cancer | Non haematological malignancy |
| C56X | Malignant neoplasm of ovary | Non haematological malignancy excluding non melonoma skin cancer | Non haematological malignancy |
| C570 | Malignant neoplasm: Fallopian tube | Non haematological malignancy excluding non melonoma skin cancer | Non haematological malignancy |
| C571 | Malignant neoplasm: Broad ligament | Non haematological malignancy excluding non melonoma skin cancer | Non haematological malignancy |
| C572 | Malignant neoplasm: Round ligament | Non haematological malignancy excluding non melonoma skin cancer | Non haematological malignancy |
| C573 | Malignant neoplasm: Parametrium | Non haematological malignancy excluding non melonoma skin cancer | Non haematological malignancy |
| C574 | Malignant neoplasm: Uterine adnexa, unspecified | Non haematological malignancy excluding non melonoma skin cancer | Non haematological malignancy |
| C577 | Malignant neoplasm: Other specified female genital organs | Non haematological malignancy excluding non melonoma skin cancer | Non haematological malignancy |
| C578 | Malignant neoplasm: Overlapping lesion of female genital organs | Non haematological malignancy excluding non melonoma skin cancer | Non haematological malignancy |
| C579 | Malignant neoplasm: Female genital organ, unspecified | Non haematological malignancy excluding non melonoma skin cancer | Non haematological malignancy |
| C58 | Malignant neoplasm of placenta | Non haematological malignancy excluding non melonoma skin cancer | Non haematological malignancy |
| C580 | Malignant neoplasm of placenta | Non haematological malignancy excluding non melonoma skin cancer | Non haematological malignancy |
| C58X | Malignant neoplasm of placenta | Non haematological malignancy excluding non melonoma skin cancer | Non haematological malignancy |
| C600 | Malignant neoplasm: Prepuce | Non haematological malignancy excluding non melonoma skin cancer | Non haematological malignancy |
| C601 | Malignant neoplasm: Glans penis | Non haematological malignancy excluding non melonoma skin cancer | Non haematological malignancy |
| C602 | Malignant neoplasm: Body of penis | Non haematological malignancy excluding non melonoma skin cancer | Non haematological malignancy |
| C608 | Malignant neoplasm: Overlapping lesion of penis | Non haematological malignancy excluding non melonoma skin cancer | Non haematological malignancy |
| C609 | Malignant neoplasm: Penis, unspecified | Non haematological malignancy excluding non melonoma skin cancer | Non haematological malignancy |
| C61 | Malignant neoplasm of prostate | Non haematological malignancy excluding non melonoma skin cancer | Non haematological malignancy |
| C610 | Malignant neoplasm of prostate | Non haematological malignancy excluding non melonoma skin cancer | Non haematological malignancy |
| C61X | Malignant neoplasm of prostate | Non haematological malignancy excluding non melonoma skin cancer | Non haematological malignancy |
| C620 | Malignant neoplasm: Undescended testis | Non haematological malignancy excluding non melonoma skin cancer | Non haematological malignancy |
| C621 | Malignant neoplasm: Descended testis | Non haematological malignancy excluding non melonoma skin cancer | Non haematological malignancy |
| C629 | Malignant neoplasm: Testis, unspecified | Non haematological malignancy excluding non melonoma skin cancer | Non haematological malignancy |
| C630 | Malignant neoplasm: Epididymis | Non haematological malignancy excluding non melonoma skin cancer | Non haematological malignancy |
| C631 | Malignant neoplasm: Spermatic cord | Non haematological malignancy excluding non melonoma skin cancer | Non haematological malignancy |
| C632 | Malignant neoplasm: Scrotum | Non haematological malignancy excluding non melonoma skin cancer | Non haematological malignancy |
| C637 | Malignant neoplasm: Other specified male genital organs | Non haematological malignancy excluding non melonoma skin cancer | Non haematological malignancy |
| C638 | Malignant neoplasm: Overlapping lesion of male genital organs | Non haematological malignancy excluding non melonoma skin cancer | Non haematological malignancy |
| C639 | Malignant neoplasm: Male genital organ, unspecified | Non haematological malignancy excluding non melonoma skin cancer | Non haematological malignancy |
| C64 | Malignant neoplasm of kidney, except renal pelvis | Non haematological malignancy excluding non melonoma skin cancer | Non haematological malignancy |
| C640 | Malignant neoplasm of kidney, except renal pelvis | Non haematological malignancy excluding non melonoma skin cancer | Non haematological malignancy |
| C64X | Malignant neoplasm of kidney, except renal pelvis | Non haematological malignancy excluding non melonoma skin cancer | Non haematological malignancy |
| C65 | Malignant neoplasm of renal pelvis | Non haematological malignancy excluding non melonoma skin cancer | Non haematological malignancy |
| C650 | Malignant neoplasm of renal pelvis | Non haematological malignancy excluding non melonoma skin cancer | Non haematological malignancy |
| C65X | Malignant neoplasm of renal pelvis | Non haematological malignancy excluding non melonoma skin cancer | Non haematological malignancy |
| C66 | Malignant neoplasm of ureter | Non haematological malignancy excluding non melonoma skin cancer | Non haematological malignancy |
| C660 | Malignant neoplasm of ureter | Non haematological malignancy excluding non melonoma skin cancer | Non haematological malignancy |
| C66X | Malignant neoplasm of ureter | Non haematological malignancy excluding non melonoma skin cancer | Non haematological malignancy |
| C670 | Malignant neoplasm: Trigone of bladder | Non haematological malignancy excluding non melonoma skin cancer | Non haematological malignancy |
| C671 | Malignant neoplasm: Dome of bladder | Non haematological malignancy excluding non melonoma skin cancer | Non haematological malignancy |
| C672 | Malignant neoplasm: Lateral wall of bladder | Non haematological malignancy excluding non melonoma skin cancer | Non haematological malignancy |
| C673 | Malignant neoplasm: Anterior wall of bladder | Non haematological malignancy excluding non melonoma skin cancer | Non haematological malignancy |
| C674 | Malignant neoplasm: Posterior wall of bladder | Non haematological malignancy excluding non melonoma skin cancer | Non haematological malignancy |
| C675 | Malignant neoplasm: Bladder neck | Non haematological malignancy excluding non melonoma skin cancer | Non haematological malignancy |
| C676 | Malignant neoplasm: Ureteric orifice | Non haematological malignancy excluding non melonoma skin cancer | Non haematological malignancy |
| C677 | Malignant neoplasm: Urachus | Non haematological malignancy excluding non melonoma skin cancer | Non haematological malignancy |
| C678 | Malignant neoplasm: Overlapping lesion of bladder | Non haematological malignancy excluding non melonoma skin cancer | Non haematological malignancy |
| C679 | Malignant neoplasm: Bladder, unspecified | Non haematological malignancy excluding non melonoma skin cancer | Non haematological malignancy |
| C680 | Malignant neoplasm: Urethra | Non haematological malignancy excluding non melonoma skin cancer | Non haematological malignancy |
| C681 | Malignant neoplasm: Paraurethral gland | Non haematological malignancy excluding non melonoma skin cancer | Non haematological malignancy |
| C688 | Malignant neoplasm: Overlapping lesion of urinary organs | Non haematological malignancy excluding non melonoma skin cancer | Non haematological malignancy |
| C689 | Malignant neoplasm: Urinary organ, unspecified | Non haematological malignancy excluding non melonoma skin cancer | Non haematological malignancy |
| C690 | Malignant neoplasm: Conjunctiva | Non haematological malignancy excluding non melonoma skin cancer | Non haematological malignancy |
| C691 | Malignant neoplasm: Cornea | Non haematological malignancy excluding non melonoma skin cancer | Non haematological malignancy |
| C692 | Malignant neoplasm: Retina | Non haematological malignancy excluding non melonoma skin cancer | Non haematological malignancy |
| C693 | Malignant neoplasm: Choroid | Non haematological malignancy excluding non melonoma skin cancer | Non haematological malignancy |
| C694 | Malignant neoplasm: Ciliary body | Non haematological malignancy excluding non melonoma skin cancer | Non haematological malignancy |
| C695 | Malignant neoplasm: Lacrimal gland and duct | Non haematological malignancy excluding non melonoma skin cancer | Non haematological malignancy |
| C696 | Malignant neoplasm: Orbit | Non haematological malignancy excluding non melonoma skin cancer | Non haematological malignancy |
| C698 | Malignant neoplasm: Overlapping lesion of eye and adnexa | Non haematological malignancy excluding non melonoma skin cancer | Non haematological malignancy |
| C699 | Malignant neoplasm: Eye, unspecified | Non haematological malignancy excluding non melonoma skin cancer | Non haematological malignancy |
| C700 | Malignant neoplasm: Cerebral meninges | Non haematological malignancy excluding non melonoma skin cancer | Non haematological malignancy |
| C701 | Malignant neoplasm: Spinal meninges | Non haematological malignancy excluding non melonoma skin cancer | Non haematological malignancy |
| C709 | Malignant neoplasm: Meninges, unspecified | Non haematological malignancy excluding non melonoma skin cancer | Non haematological malignancy |
| C710 | Malignant neoplasm: Cerebrum, except lobes and ventricles | Non haematological malignancy excluding non melonoma skin cancer | Non haematological malignancy |
| C711 | Malignant neoplasm: Frontal lobe | Non haematological malignancy excluding non melonoma skin cancer | Non haematological malignancy |
| C712 | Malignant neoplasm: Temporal lobe | Non haematological malignancy excluding non melonoma skin cancer | Non haematological malignancy |
| C713 | Malignant neoplasm: Parietal lobe | Non haematological malignancy excluding non melonoma skin cancer | Non haematological malignancy |
| C714 | Malignant neoplasm: Occipital lobe | Non haematological malignancy excluding non melonoma skin cancer | Non haematological malignancy |
| C715 | Malignant neoplasm: Cerebral ventricle | Non haematological malignancy excluding non melonoma skin cancer | Non haematological malignancy |
| C716 | Malignant neoplasm: Cerebellum | Non haematological malignancy excluding non melonoma skin cancer | Non haematological malignancy |
| C717 | Malignant neoplasm: Brain stem | Non haematological malignancy excluding non melonoma skin cancer | Non haematological malignancy |
| C718 | Malignant neoplasm: Overlapping lesion of brain | Non haematological malignancy excluding non melonoma skin cancer | Non haematological malignancy |
| C719 | Malignant neoplasm: Brain, unspecified | Non haematological malignancy excluding non melonoma skin cancer | Non haematological malignancy |
| C720 | Malignant neoplasm: Spinal cord | Non haematological malignancy excluding non melonoma skin cancer | Non haematological malignancy |
| C721 | Malignant neoplasm: Cauda equina | Non haematological malignancy excluding non melonoma skin cancer | Non haematological malignancy |
| C722 | Malignant neoplasm: Olfactory nerve | Non haematological malignancy excluding non melonoma skin cancer | Non haematological malignancy |
| C723 | Malignant neoplasm: Optic nerve | Non haematological malignancy excluding non melonoma skin cancer | Non haematological malignancy |
| C724 | Malignant neoplasm: Acoustic nerve | Non haematological malignancy excluding non melonoma skin cancer | Non haematological malignancy |
| C725 | Malignant neoplasm: Other and unspecified cranial nerves | Non haematological malignancy excluding non melonoma skin cancer | Non haematological malignancy |
| C728 | Malignant neoplasm: Overlapping lesion of brain and other parts of central nervous system | Non haematological malignancy excluding non melonoma skin cancer | Non haematological malignancy |
| C729 | Malignant neoplasm: Central nervous system, unspecified | Non haematological malignancy excluding non melonoma skin cancer | Non haematological malignancy |
| C73 | Malignant neoplasm of thyroid gland | Non haematological malignancy excluding non melonoma skin cancer | Non haematological malignancy |
| C730 | Malignant neoplasm of thyroid gland | Non haematological malignancy excluding non melonoma skin cancer | Non haematological malignancy |
| C73X | Malignant neoplasm of thyroid gland | Non haematological malignancy excluding non melonoma skin cancer | Non haematological malignancy |
| C740 | Malignant neoplasm: Cortex of adrenal gland | Non haematological malignancy excluding non melonoma skin cancer | Non haematological malignancy |
| C741 | Malignant neoplasm: Medulla of adrenal gland | Non haematological malignancy excluding non melonoma skin cancer | Non haematological malignancy |
| C749 | Malignant neoplasm: Adrenal gland, unspecified | Non haematological malignancy excluding non melonoma skin cancer | Non haematological malignancy |
| C750 | Malignant neoplasm: Parathyroid gland | Non haematological malignancy excluding non melonoma skin cancer | Non haematological malignancy |
| C751 | Malignant neoplasm: Pituitary gland | Non haematological malignancy excluding non melonoma skin cancer | Non haematological malignancy |
| C752 | Malignant neoplasm: Craniopharyngeal duct | Non haematological malignancy excluding non melonoma skin cancer | Non haematological malignancy |
| C753 | Malignant neoplasm: Pineal gland | Non haematological malignancy excluding non melonoma skin cancer | Non haematological malignancy |
| C754 | Malignant neoplasm: Carotid body | Non haematological malignancy excluding non melonoma skin cancer | Non haematological malignancy |
| C755 | Malignant neoplasm: Aortic body and other paraganglia | Non haematological malignancy excluding non melonoma skin cancer | Non haematological malignancy |
| C758 | Malignant neoplasm: Pluriglandular involvement, unspecified | Non haematological malignancy excluding non melonoma skin cancer | Non haematological malignancy |
| C759 | Malignant neoplasm: Endocrine gland, unspecified | Non haematological malignancy excluding non melonoma skin cancer | Non haematological malignancy |
| C760 | Malignant neoplasm of other and ill-defined sites: Head, face and neck | Non haematological malignancy excluding non melonoma skin cancer | Non haematological malignancy |
| C761 | Malignant neoplasm of other and ill-defined sites: Thorax | Non haematological malignancy excluding non melonoma skin cancer | Non haematological malignancy |
| C762 | Malignant neoplasm of other and ill-defined sites: Abdomen | Non haematological malignancy excluding non melonoma skin cancer | Non haematological malignancy |
| C763 | Malignant neoplasm of other and ill-defined sites: Pelvis | Non haematological malignancy excluding non melonoma skin cancer | Non haematological malignancy |
| C764 | Malignant neoplasm of other and ill-defined sites: Upper limb | Non haematological malignancy excluding non melonoma skin cancer | Non haematological malignancy |
| C765 | Malignant neoplasm of other and ill-defined sites: Lower limb | Non haematological malignancy excluding non melonoma skin cancer | Non haematological malignancy |
| C767 | Malignant neoplasm of other and ill-defined sites: Other ill-defined sites | Non haematological malignancy excluding non melonoma skin cancer | Non haematological malignancy |
| C768 | Malignant neoplasm of other and ill-defined sites: Overlapping lesion of other and ill-defined sites | Non haematological malignancy excluding non melonoma skin cancer | Non haematological malignancy |
| C770 | Secondary and unspecified malignant neoplasm: Lymph nodes of head, face and neck | Non haematological malignancy excluding non melonoma skin cancer | Non haematological malignancy |
| C771 | Secondary and unspecified malignant neoplasm: Intrathoracic lymph nodes | Non haematological malignancy excluding non melonoma skin cancer | Non haematological malignancy |
| C772 | Secondary and unspecified malignant neoplasm: Intra-abdominal lymph nodes | Non haematological malignancy excluding non melonoma skin cancer | Non haematological malignancy |
| C773 | Secondary and unspecified malignant neoplasm: Axillary and upper limb lymph nodes | Non haematological malignancy excluding non melonoma skin cancer | Non haematological malignancy |
| C774 | Secondary and unspecified malignant neoplasm: Inguinal and lower limb lymph nodes | Non haematological malignancy excluding non melonoma skin cancer | Non haematological malignancy |
| C775 | Secondary and unspecified malignant neoplasm: Intrapelvic lymph nodes | Non haematological malignancy excluding non melonoma skin cancer | Non haematological malignancy |
| C778 | Secondary and unspecified malignant neoplasm: Lymph nodes of multiple regions | Non haematological malignancy excluding non melonoma skin cancer | Non haematological malignancy |
| C779 | Secondary and unspecified malignant neoplasm: Lymph node, unspecified | Non haematological malignancy excluding non melonoma skin cancer | Non haematological malignancy |
| C780 | Secondary malignant neoplasm of lung | Non haematological malignancy excluding non melonoma skin cancer | Non haematological malignancy |
| C781 | Secondary malignant neoplasm of mediastinum | Non haematological malignancy excluding non melonoma skin cancer | Non haematological malignancy |
| C782 | Secondary malignant neoplasm of pleura | Non haematological malignancy excluding non melonoma skin cancer | Non haematological malignancy |
| C783 | Secondary malignant neoplasm of other and unspecified respiratory organs | Non haematological malignancy excluding non melonoma skin cancer | Non haematological malignancy |
| C784 | Secondary malignant neoplasm of small intestine | Non haematological malignancy excluding non melonoma skin cancer | Non haematological malignancy |
| C785 | Secondary malignant neoplasm of large intestine and rectum | Non haematological malignancy excluding non melonoma skin cancer | Non haematological malignancy |
| C786 | Secondary malignant neoplasm of retroperitoneum and peritoneum | Non haematological malignancy excluding non melonoma skin cancer | Non haematological malignancy |
| C787 | Secondary malignant neoplasm of liver and intrahepatic bile duct | Non haematological malignancy excluding non melonoma skin cancer | Non haematological malignancy |
| C788 | Secondary malignant neoplasm of other and unspecified digestive organs | Non haematological malignancy excluding non melonoma skin cancer | Non haematological malignancy |
| C790 | Secondary malignant neoplasm of kidney and renal pelvis | Non haematological malignancy excluding non melonoma skin cancer | Non haematological malignancy |
| C791 | Secondary malignant neoplasm of bladder and other and unspecified urinary organs | Non haematological malignancy excluding non melonoma skin cancer | Non haematological malignancy |
| C792 | Secondary malignant neoplasm of skin | Non haematological malignancy excluding non melonoma skin cancer | Non haematological malignancy |
| C793 | Secondary malignant neoplasm of brain and cerebral meninges | Non haematological malignancy excluding non melonoma skin cancer | Non haematological malignancy |
| C794 | Secondary malignant neoplasm of other and unspecified parts of nervous system | Non haematological malignancy excluding non melonoma skin cancer | Non haematological malignancy |
| C795 | Secondary malignant neoplasm of bone and bone marrow | Non haematological malignancy excluding non melonoma skin cancer | Non haematological malignancy |
| C796 | Secondary malignant neoplasm of ovary | Non haematological malignancy excluding non melonoma skin cancer | Non haematological malignancy |
| C797 | Secondary malignant neoplasm of adrenal gland | Non haematological malignancy excluding non melonoma skin cancer | Non haematological malignancy |
| C798 | Secondary malignant neoplasm of other specified sites | Non haematological malignancy excluding non melonoma skin cancer | Non haematological malignancy |
| C80 | Malignant neoplasm without specification of site | Non haematological malignancy excluding non melonoma skin cancer | Non haematological malignancy |
| C800 | Malignant neoplasm, primary site unknown, so stated | Non haematological malignancy excluding non melonoma skin cancer | Non haematological malignancy |
| C80X | Malignant neoplasm without specification of site | Non haematological malignancy excluding non melonoma skin cancer | Non haematological malignancy |
| C81 | Hodgkin lymphoma | Hodgkin lymphoma | Haematological malignancy |
| C810 | Nodular lymphocyte predominant Hodgkin lymphoma | B-cell lymphoma | Haematological malignancy |
| C811 | Nodular sclerosis (classical) Hodgkin lymphoma | Hodgkin lymphoma | Haematological malignancy |
| C812 | Mixed cellularity (classical) Hodgkin lymphoma | Hodgkin lymphoma | Haematological malignancy |
| C813 | Lymphocyte depleted (classical) Hodgkin lymphoma | Hodgkin lymphoma | Haematological malignancy |
| C814 | Lymphocyte-rich (classical) Hodgkin lymphoma | Hodgkin lymphoma | Haematological malignancy |
| C817 | Other (classical) Hodgkin lymphoma | Hodgkin lymphoma | Haematological malignancy |
| C819 | Hodgkin lymphoma, unspecified | Hodgkin lymphoma | Haematological malignancy |
| C81X | Hodgkin lymphoma | Hodgkin lymphoma | Haematological malignancy |
| C82 | Follicular lymphoma | B-cell lymphoma | Haematological malignancy |
| C820 | Follicular lymphoma grade I | B-cell lymphoma | Haematological malignancy |
| C821 | Follicular lymphoma grade II | B-cell lymphoma | Haematological malignancy |
| C822 | Follicular lymphoma grade III, unspecified | B-cell lymphoma | Haematological malignancy |
| C823 | Follicular lymphoma grade IIIa | B-cell lymphoma | Haematological malignancy |
| C824 | Follicular lymphoma grade IIIb | B-cell lymphoma | Haematological malignancy |
| C825 | Diffuse follicle centre lymphoma | B-cell lymphoma | Haematological malignancy |
| C826 | Cutaneous follicle centre lymphoma | B-cell lymphoma | Haematological malignancy |
| C827 | Other types of follicular lymphoma | B-cell lymphoma | Haematological malignancy |
| C829 | Follicular lymphoma, unspecified | B-cell lymphoma | Haematological malignancy |
| C82X | Follicular lymphoma | B-cell lymphoma | Haematological malignancy |
| C83 | Non-follicular lymphoma | B-cell lymphoma | Haematological malignancy |
| C830 | Small cell B-cell lymphoma | B-cell lymphoma | Haematological malignancy |
| C831 | Mantle cell lymphoma | B-cell lymphoma | Haematological malignancy |
| C833 | Diffuse large B-cell lymphoma | B-cell lymphoma or Non haematological malignancy excluding non melonoma skin cancer depending on associated morphology code | Haematological malignancy or Non haematological malignancy depending on associated morphology code |
| C835 | Lymphoblastic (diffuse) lymphoma | Leukaemia | Haematological malignancy |
| C837 | Burkitt lymphoma | B-cell lymphoma | Haematological malignancy |
| C838 | Other non-follicular lymphoma | B-cell lymphoma or Lymphoma NOS depending on associated morphology code | Haematological malignancy |
| C839 | Non-follicular (diffuse) lymphoma, unspecified | B-cell lymphoma | Haematological malignancy |
| C83X | Non-follicular lymphoma | B-cell lymphoma | Haematological malignancy |
| C84 | Mature T/NK-cell lymphomas | T-cell lymphoma | Haematological malignancy |
| C840 | Mycosis fungoides | T-cell lymphoma | Haematological malignancy |
| C841 | Sézary disease | T-cell lymphoma | Haematological malignancy |
| C844 | Peripheral T-cell lymphoma, not elsewhere classified | T-cell lymphoma | Haematological malignancy |
| C845 | Other mature T/NK-cell lymphomas | T-cell lymphoma | Haematological malignancy |
| C846 | Anaplastic large cell lymphoma, ALK-positive | T-cell lymphoma | Haematological malignancy |
| C847 | Anaplastic large cell lymphoma, ALK-negative | T-cell lymphoma | Haematological malignancy |
| C848 | Cutaneous T-cell lymphoma, unspecified | T-cell lymphoma | Haematological malignancy |
| C849 | Mature T/NK-cell lymphoma, unspecified | T-cell lymphoma | Haematological malignancy |
| C84X | Mature T/NK-cell lymphomas | Lymphoma NOS | Haematological malignancy |
| C85 | Other and unspecified types of non-Hodgkin lymphoma | Lymphoma NOS | Haematological malignancy |
| C850 | Other and unspecified types of non-Hodgkin lymphoma | T-cell lymphoma or B-cell lymphoma or Lymphoma NOS depending on associated morphology code | Haematological malignancy |
| C851 | B-cell lymphoma, unspecified | B-cell lymphoma | Haematological malignancy |
| C852 | Mediastinal (thymic) large B-cell lymphoma | B-cell lymphoma | Haematological malignancy |
| C857 | Other specified types of non-Hodgkin lymphoma | T-cell lymphoma or B-cell lymphoma or Lymphoma NOS depending on associated morphology code | Haematological malignancy |
| C859 | Non-Hodgkin lymphoma, unspecified | Lymphoma NOS | Haematological malignancy |
| C85X | Other and unspecified types of non-Hodgkin lymphoma | T-cell lymphoma or B-cell lymphoma or Lymphoma NOS depending on associated morphology code | Haematological malignancy |
| C86 | Other specified types of T/NK-cell lymphoma | Lymphoma NOS | Haematological malignancy |
| C860 | Extranodal NK/T-cell lymphoma, nasal type | T-cell lymphoma | Haematological malignancy |
| C861 | Hepatosplenic T-cell lymphoma | T-cell lymphoma | Haematological malignancy |
| C862 | Enteropathy-type (intestinal) T-cell lymphoma | T-cell lymphoma | Haematological malignancy |
| C863 | Subcutaneous panniculitis-like T-cell lymphoma | T-cell lymphoma | Haematological malignancy |
| C864 | Blastic NK-cell lymphoma | Lymphoma NOS | Haematological malignancy |
| C865 | Angioimmunoblastic T-cell lymphoma | T-cell lymphoma | Haematological malignancy |
| C866 | Primary cutaneous CD30-positive T-cell proliferations | T-cell lymphoma | Haematological malignancy |
| C86X | Other specified types of T/NK-cell lymphoma | Lymphoma NOS | Haematological malignancy |
| C88 | Malignant immunoproliferative diseases | T-cell lymphoma or B-cell lymphoma depending on associated morphology code | Haematological malignancy |
| C880 | Waldenström macroglobulinaemia | B-cell lymphoma | Haematological malignancy |
| C882 | Other heavy chain disease | B-cell lymphoma | Haematological malignancy |
| C883 | Immunoproliferative small intestinal disease | B-cell lymphoma | Haematological malignancy |
| C884 | Extranodal marginal zone B-cell lymphoma of mucosa-associated lymphoid tissue [MALT-lymphoma] | B-cell lymphoma | Haematological malignancy |
| C887 | Other malignant immunoproliferative diseases | Lymphoma NOS | Haematological malignancy |
| C889 | Malignant immunoproliferative disease, unspecified | Lymphoma NOS | Haematological malignancy |
| C88X | Malignant immunoproliferative diseases | T-cell lymphoma or B-cell lymphoma depending on associated morphology code | Haematological malignancy |
| C90 | Multiple myeloma and malignant plasma cell neoplasms | Other haematological histiocytic / Myelodysplastic / Malignancy / Unspecified | Haematological malignancy |
| C900 | Multiple myeloma | Other haematological histiocytic / Myelodysplastic / Malignancy / Unspecified | Haematological malignancy |
| C901 | Plasma cell leukaemia | Leukaemia | Haematological malignancy |
| C902 | Extramedullary plasmacytoma | Other haematological histiocytic / Myelodysplastic / Malignancy / Unspecified | Haematological malignancy |
| C903 | Solitary plasmacytoma | Other haematological histiocytic / Myelodysplastic / Malignancy / Unspecified | Haematological malignancy |
| C90X | Multiple myeloma and malignant plasma cell neoplasms | Other haematological histiocytic / Myelodysplastic / Malignancy / Unspecified | Haematological malignancy |
| C91 | Lymphoid leukaemia | Leukaemia | Haematological malignancy |
| C910 | Acute lymphoblastic leukaemia [ALL] | Leukaemia | Haematological malignancy |
| C911 | Chronic lymphocytic leukaemia of B-cell type | Leukaemia | Haematological malignancy |
| C913 | Prolymphocytic leukaemia of B-cell type | Leukaemia or Other haematological histiocytic / Myelodysplastic / Malignancy / Unspecified depending on associated morphology code | Haematological malignancy |
| C914 | Hairy-cell leukaemia | B-cell lymphoma | Haematological malignancy |
| C915 | Adult T-cell lymphoma/leukaemia [HTLV-1-associated] | T-cell lymphoma | Haematological malignancy |
| C916 | Prolymphocytic leukaemia of T-cell type | Leukaemia | Haematological malignancy |
| C917 | Other lymphoid leukaemia | B-cell lymphoma | Haematological malignancy |
| C918 | Mature B-cell leukaemia Burkitt-type | B-cell lymphoma | Haematological malignancy |
| C919 | Lymphoid leukaemia, unspecified | Leukaemia or T-cell lymphoma depending on associated morphology code | Haematological malignancy |
| C91X | Lymphoid Leukaemia | Leukaemia | Haematological malignancy |
| C92 | Myeloid leukaemia | Leukaemia or Other haematological histiocytic / Myelodysplastic / Malignancy / Unspecified | Haematological malignancy |
| C920 | Acute myeloblastic leukaemia [AML] | Leukaemia | Haematological malignancy |
| C921 | Chronic myeloid leukaemia [CML], BCR/ABL-positive | Leukaemia | Haematological malignancy |
| C922 | Atypical chronic myeloid leukaemia, BCR/ABL-negative | Leukaemia | Haematological malignancy |
| C923 | Myeloid sarcoma | Leukaemia | Haematological malignancy |
| C924 | Acute promyelocytic leukaemia [PML] | Leukaemia | Haematological malignancy |
| C925 | Acute myelomonocytic leukaemia | Leukaemia | Haematological malignancy |
| C926 | Acute myeloid leukaemia with 11q23-abnormality | Leukaemia | Haematological malignancy |
| C927 | Other myeloid leukaemia | Leukaemia | Haematological malignancy |
| C928 | Acute myeloid leukaemia with multilineage dysplasia | Leukaemia | Haematological malignancy |
| C929 | Myeloid leukaemia, unspecified | Leukaemia | Haematological malignancy |
| C92X | Myeloid Leukaemia | Leukaemia | Haematological malignancy |
| C93 | Monocytic leukaemia | Leukaemia | Haematological malignancy |
| C930 | Acute monoblastic/monocytic leukaemia | Leukaemia | Haematological malignancy |
| C930 | Acute monoblastic/monocytic leukaemia | Leukaemia | Haematological malignancy |
| C931 | Chronic myelomonocytic leukaemia | Leukaemia | Haematological malignancy |
| C933 | Juvenile myelomonocytic leukaemia | Leukaemia | Haematological malignancy |
| C937 | Other monocytic leukaemia | Leukaemia | Haematological malignancy |
| C939 | Monocytic leukaemia, unspecified | Leukaemia | Haematological malignancy |
| C93X | Monocytic Leukaemia | Leukaemia | Haematological malignancy |
| C94 | Other leukaemias of specified cell type | Leukaemia | Haematological malignancy |
| C940 | Acute erythroid leukaemia | Leukaemia | Haematological malignancy |
| C942 | Acute megakaryoblastic leukaemia | Leukaemia | Haematological malignancy |
| C943 | Mast cell leukaemia | Leukaemia | Haematological malignancy |
| C944 | Acute panmyelosis with myelofibrosis | Other haematological histiocytic / Myelodysplastic / Malignancy / Unspecified | Haematological malignancy |
| C946 | Myelodysplastic and myeloproliferative disease, not elsewhere classified | Other haematological histiocytic / Myelodysplastic / Malignancy / Unspecified | Haematological malignancy |
| C947 | Other specified leukaemias | Leukaemia | Haematological malignancy |
| C94X | Other Leukaemias of specified cell type | Leukaemia | Haematological malignancy |
| C95 | Leukaemia of unspecified cell type | Leukaemia | Haematological malignancy |
| C950 | Acute leukaemia of unspecified cell type | Lymphoma NOS or Leukaemia depending on associated morphology code | Haematological malignancy |
| C951 | Chronic leukaemia of unspecified cell type | Leukaemia | Haematological malignancy |
| C957 | Other leukaemia of unspecified cell type | Leukaemia | Haematological malignancy |
| C959 | Leukaemia, unspecified | Other haematological histiocytic / Myelodysplastic / Malignancy / Unspecified | Haematological malignancy |
| C95X | Leukaemia of unspecified cell type | Leukaemia | Haematological malignancy |
| C96 | Other and unspecified malignant neoplasms of lymphoid, haematopoietic and related tissue | Other haematological histiocytic / Myelodysplastic / Malignancy / Unspecified | Haematological malignancy |
| C960 | Multifocal and multisystemic (disseminated) Langerhans-cell histiocytosis [Letterer-Siwe disease] | Other haematological histiocytic / Myelodysplastic / Malignancy / Unspecified | Haematological malignancy |
| C962 | Malignant mast cell tumour | Other haematological histiocytic / Myelodysplastic / Malignancy / Unspecified | Haematological malignancy |
| C964 | Sarcoma of dendritic cells (accessory cells) | Other haematological histiocytic / Myelodysplastic / Malignancy / Unspecified | Haematological malignancy |
| C965 | Multifocal and unisystemic Langerhans-cell histiocytosis | Other haematological histiocytic / Myelodysplastic / Malignancy / Unspecified | Haematological malignancy |
| C966 | Unifocal Langerhans-cell histiocytosis | Other haematological histiocytic / Myelodysplastic / Malignancy / Unspecified | Haematological malignancy |
| C967 | Other specified malignant neoplasms of lymphoid, haematopoietic and related tissue | Other haematological histiocytic / Myelodysplastic / Malignancy / Unspecified | Haematological malignancy |
| C968 | Histiocytic sarcoma | Other haematological histiocytic / Myelodysplastic / Malignancy / Unspecified | Haematological malignancy |
| C969 | Malignant neoplasm of lymphoid, haematopoietic and related tissue, unspecified | Other haematological histiocytic / Myelodysplastic / Malignancy / Unspecified | Haematological malignancy |
| C96X | Other and unspecified malignant neoplasms of lymphoid, haematopoietic and related tissue | Other haematological histiocytic / Myelodysplastic / Malignancy / Unspecified | Haematological malignancy |
| D020 | Carcinoma in situ: Larynx | Non haematological malignancy excluding non melonoma skin cancer | Non haematological malignancy |
| D033 | Melanoma in situ of other and unspecified parts of face | Non haematological malignancy excluding non melonoma skin cancer | Non haematological malignancy |
| D050 | Lobular carcinoma in situ | Non haematological malignancy excluding non melonoma skin cancer | Non haematological malignancy |
| D051 | Intraductal carcinoma in situ | Non haematological malignancy excluding non melonoma skin cancer | Non haematological malignancy |
| D075 | Carcinoma in situ: Prostate | Non haematological malignancy excluding non melonoma skin cancer | Non haematological malignancy |
| D090 | Carcinoma in situ: Bladder | Non haematological malignancy excluding non melonoma skin cancer | Non haematological malignancy |
| D410 | Neoplasm of uncertain or unknown behaviour: Kidney | Non haematological malignancy excluding non melonoma skin cancer | Non haematological malignancy |
| D439 | Neoplasm of uncertain or unknown behaviour: Central nervous system, unspecified | Other haematological histiocytic / Myelodysplastic / Malignancy / Unspecified | Haematological malignancy |
| D45 | Polycythaemia vera | Other haematological histiocytic / Myelodysplastic / Malignancy / Unspecified | Haematological malignancy |
| D461 | Refractory anaemia with ring sideroblasts | Other haematological histiocytic / Myelodysplastic / Malignancy / Unspecified | Haematological malignancy |
| D462 | Refractory anaemia with excess of blasts [RAEB] | Other haematological histiocytic / Myelodysplastic / Malignancy / Unspecified | Haematological malignancy |
| D464 | Refractory anaemia, unspecified | Other haematological histiocytic / Myelodysplastic / Malignancy / Unspecified | Haematological malignancy |
| D469 | Myelodysplastic syndrome, unspecified | Other haematological histiocytic / Myelodysplastic / Malignancy / Unspecified | Haematological malignancy |
| D470 | Histiocytic and mast cell tumours of uncertain and unknown behaviour | Other haematological histiocytic / Myelodysplastic / Malignancy / Unspecified | Haematological malignancy |
| D471 | Chronic myeloproliferative disease | Other haematological histiocytic / Myelodysplastic / Malignancy / Unspecified | Haematological malignancy |
| D473 | Essential (haemorrhagic) thrombocythaemia | Other haematological histiocytic / Myelodysplastic / Malignancy / Unspecified | Haematological malignancy |
| D477 | Other specified neoplasms of uncertain or unknown behaviour of lymphoid, haematopoietic and related tissue | Other haematological histiocytic / Myelodysplastic / Malignancy / Unspecified | Haematological malignancy |
| D479 | Neoplasm of uncertain or unknown behaviour of lymphoid, haematopoietic and related tissue, unspecified | Other haematological histiocytic / Myelodysplastic / Malignancy / Unspecified or B-cell lymphoma depending on associated morphology code | Haematological malignancy |
| D485 | Neoplasm of uncertain or unknown behaviour: Skin | Other haematological histiocytic / Myelodysplastic / Malignancy / Unspecified | Haematological malignancy |
| D489 | Neoplasm of uncertain or unknown behaviour: Neoplasm of uncertain or unknown behaviour, unspecified | Non haematological malignancy excluding non melonoma skin cancer | Non haematological malignancy |
