## Supplementary Table 4 for "Temporal trends in the incidence of haemophagocytic lymphohistiocytosis: a nationwide cohort study from England 2003-2018"

Table S4. Mutually adjusted Incidence Rate Ratios (IRR) and 95% confidence intervals for HLH.

| **Characteristic** | **IRR***^1^* | **95% CI***^1^* | **p-value** |
| --- | --- | --- | --- |
| Year of diagnosis |  |  |  |
| 2003 | — | — |  |
| 2004 | 0.94 | 0.62, 1.40 | 0.8 |
| 2005 | 0.82 | 0.55, 1.22 | 0.3 |
| 2006 | 1.01 | 0.70, 1.48 | >0.9 |
| 2007 | 1.15 | 0.80, 1.66 | 0.5 |
| 2008 | 1.14 | 0.79, 1.64 | 0.5 |
| 2009 | 1.37 | 0.97, 1.96 | 0.075 |
| 2010 | 1.52 | 1.08, 2.15 | 0.017 |
| 2011 | 1.66 | 1.19, 2.34 | 0.003 |
| 2012 | 2.08 | 1.51, 2.89 | <0.001 |
| 2013 | 2.09 | 1.52, 2.91 | <0.001 |
| 2014 | 2.39 | 1.76, 3.31 | <0.001 |
| 2015 | 2.39 | 1.75, 3.30 | <0.001 |
| 2016 | 2.35 | 1.73, 3.25 | <0.001 |
| 2017 | 3.21 | 2.39, 4.40 | <0.001 |
| 2018 | 3.88 | 2.91, 5.28 | <0.001 |
| Age group |  |  |  |
| 0-4 | — | — |  |
| 5-14 | 0.31 | 0.26, 0.37 | <0.001 |
| 15-54 | 0.20 | 0.18, 0.24 | <0.001 |
| 55+ | 0.39 | 0.34, 0.45 | <0.001 |
| Sex |  |  |  |
| Female | — | — |  |
| Male | 1.32 | 1.20, 1.46 | <0.001 |
| *^1^* IRR = Incidence Rate Ratio, CI = Confidence Interval | | | |
